# Microbiota effects and predictors of *Lactobacillus crispatu*s colonization after treatment with a vaginal live biotherapeutic: results from a randomized, double-blinded, placebo-controlled trial

**DOI:** 10.1101/2025.08.18.25333897

**Authors:** Seth M. Bloom, Laura Symul, Joseph Elsherbini, Jiawu Xu, Salina Hussain, Johnathan Shih, Ashley Sango, Caroline M. Mitchell, Anke Hemmerling, Thomas P. Parks, Aditi Kannan, Fatima A. Hussain, Craig R. Cohen, Susan P. Holmes, Douglas S. Kwon

## Abstract

Bacterial vaginosis (BV) affects >25% of women worldwide and often recurs after standard-of-care metronidazole (MTZ) treatment. LACTIN-V, a live biotherapeutic product (LBP) containing *Lactobacillus crispatus* strain CTV-05, reduced recurrent BV in a Phase 2b clinical trial, but efficacy was incomplete. We characterized microbiota and immune effects and correlates of treatment success in trial samples. By week 12, *L. crispatus*-dominant microbiota was achieved in 30% of LBP recipients compared to 9% of placebo (benefit ratio: 3.31; p<0.005). This effect was mostly due to CTV-05, but native *L. crispatus* strains were also present and increased over time. Inflammatory cytokines decreased in both arms after MTZ, but returned to baseline in placebo recipients. *L. crispatus* colonization was associated with pre-MTZ microbiota, baseline cytokine profiles, post-MTZ bacterial load, and clinical and behavioral variables. These findings elucidate LBP microbiota effects and identify predictors of treatment success, informing improved intervention strategies to advance women’s health.

## Introduction

Bacterial vaginosis (BV), a vaginal syndrome characterized by *Lactobacillus*-deficient vaginal microbiota, affects 23-29% of women of reproductive age globally, with an estimated annual economic burden of $4.8 billion worldwide (Peebles et al., 2019; Workowski et al., 2021). BV symptoms include vaginal discharge, malodor, pain, and itching, leading to significant impairments in quality of life, self-esteem, and sexual function (Bilardi et al., 2013; Brusselmans et al., 2023). In addition, BV is linked to mucosal inflammation and increased risk for numerous adverse health outcomes, such as HIV acquisition, sexually transmitted infections, preterm birth, human papillomavirus (HPV) infection, and cervical cancer (Anahtar et al., 2015; Atashili et al., 2008; Brusselaers et al., 2019; Gosmann et al., 2017; Hillier et al., 1988; Leitich and Kiss, 2007). While BV occurs globally, it disproportionately affects those with lower socioeconomic status and members of racial or ethnic minority groups across diverse settings (Allsworth and Peipert, 2007; Kenyon et al., 2013; Marconi et al., 2015; Peebles et al., 2019). Development of effective BV treatments is therefore a key objective to improve women’s health worldwide (Bradshaw and Sobel, 2016).

Current first-line therapy for BV consists of oral or intra-vaginal antibiotics such as metronidazole (MTZ), which target many species within the diverse anaerobic bacterial communities characteristic of BV (Bradshaw and Sobel, 2016; Workowski et al., 2021). In most cases, MTZ reduces abundance of BV-associated anaerobes and leads to emergence of bacterial communities dominated by *Lactobacillus* species (which are intrinsically MTZ-resistant) (Bradshaw and Brotman, 2015), but BV frequently recurs after treatment. An Australian study of women with symptomatic BV found that 58% experienced recurrent BV (rBV) within 1 year of MTZ therapy, while a US trial that enrolled women with symptomatic BV and a prior history of post-treatment recurrence found that >75-80% experienced rBV within 16 weeks of treatment (Bradshaw et al., 2006; Schwebke et al., 2021). It is hypothesized that the incomplete efficacy of MTZ often results from a failure to eradicate BV-associated bacterial communities and/or from their post-MTZ replacement with *Lactobacillus* species prone to reverting to BV (Bradshaw and Brotman, 2015). Studies have shown that *Lactobacillus iners* is associated with an increased risk of transition to BV-like states (DiGiulio et al., 2015; Munoz et al., 2021; Tamarelle et al., 2022) and with higher rates of adverse health outcomes (Colbert et al., 2023; Gosmann et al., 2017; Kindinger et al., 2017; Norenhag et al., 2020; van Houdt et al., 2018). MTZ treatment frequently results in establishment of vaginal microbiota dominated by *L. iners* rather than *L. crispatus* (Ferris et al., 2007; Joag et al., 2019; Mitchell et al., 2012; Ravel et al., 2013; Srinivasan et al., 2010; Verwijs et al., 2020), supporting the hypothesis that BV therapies which preferentially promote *L. crispatus* over *L. iners* may improve treatment outcomes and promote vaginal health (Bradshaw and Brotman, 2015; Nilsen et al., 2020; Zhu et al., 2024).

New strategies to enhance *L. crispatus* colonization during BV treatment are in various stages of development, including vaginal microbiome transplants, adjunctive therapies that selectively inhibit *L. iners* and/or promote *L. crispatus* growth, and *L. crispatus*-containing live biotherapeutic products (LBPs) (Bloom et al., 2022; Chetty et al., 2025; Cohen et al., 2020; Lev-Sagie et al., 2019; Nilsen et al., 2020; Ravel et al., 2025; Yockey et al., 2022; Zhu et al., 2024). The most advanced LBP in development is LACTIN-V, a vaginally-administered LBP containing the *L. crispatus* strain CTV-05, and the only LBP tested in large-scale clinical trials (Cohen et al., 2020; Hemmerling et al., 2009). Cohen and colleagues recently reported results of a phase 2b multi-center, randomized, placebo-controlled, double-blinded study showing that participants who received 11 weeks of intravaginal LACTIN-V after a 5-day course of intravaginal MTZ developed rBV at lower rates than placebo recipients (Cohen et al., 2020). Benefits from LACTIN-V were observed at both week 12 (approximately 1 week after completing therapy; relative risk of rBV = 0.66) and week 24 (approximately 13 weeks after completing therapy; relative risk of rBV = 0.73). However, rBV rates remained high even in the LBP arm, with 30% of recipients experiencing rBV by week 12 and 39% experiencing recurrence by week 24 in the intention-to-treat analysis. In prior reports, rBV was characterized using clinical measures, but comprehensive molecular analysis of vaginal microbiota composition and factors associated with colonization were not assessed.

Here we characterize the effects of LACTIN-V on vaginal microbiota composition and identify correlates of successful colonization by analyzing vaginal microbiota composition, bacterial strain dynamics, cytokine profiles, and demographic/behavioral parameters of LACTIN-V phase 2b trial participants (Cohen et al., 2020). We find that the reduction in rBV among LBP recipients corresponded to >3-fold higher rates of *L. crispatus*-dominant colonization, but only 30% of recipients achieved *L. crispatus-* dominant vaginal microbiota at week 12, consistent with the observed incomplete clinical efficacy. Microbiota trajectory and strain dynamics revealed CTV-05 accounted for the majority of *L. crispatus* colonization among LBP recipients, though endogenous strains increased over time and sometimes displaced CTV-05. We further describe the effects of LBP treatment and microbiota composition on vaginal immune markers, and show that pre-MTZ microbiota composition, baseline inflammatory cytokine profiles, post-MTZ total bacterial load, and selected clinical/behavioral variables are associated with LBP treatment success. These results show how an *L. crispatus* LBP alters vaginal microbiota composition to improve BV treatment efficacy while identifying factors linked to LBP success that can guide development of future therapies to promote more optimal vaginal microbiota communities and improved women’s health outcomes.

## Results

### Study design and participant characteristics

Women aged 18 to 45 years with BV completed five days of standard intravaginal MTZ therapy and were then randomized 2:1 to receive intravaginal LACTIN-V (Osel, Inc.) or placebo as previously described (Cohen et al., 2020). Enrollment occurred at four U.S. sites. LACTIN-V is a powder formulation containing the live *L. crispatus* strain CTV-05, administered via prefilled, single-use vaginal applicators at a dose of 2×10^9^ colony-forming units (CFU). The placebo contained the same inactive ingredients without bacteria and was visually indistinguishable. BV diagnosed at the baseline “pre-MTZ” screening visit required at least three of four Amsel criteria and a Nugent score of ≥4 (Amsel et al., 1983; Nugent et al., 1991). Participants received five days of intravaginal MTZ gel within 30 days of diagnosis, and then were seen at a “post-MTZ” visit within 48 hours of completing antibiotics for randomization to LBP or placebo. The first treatment dose was clinician-administered at the post-MTZ visit, then participants self-administered doses daily for four consecutive days, followed by twice weekly dosing for 10 weeks (ending at week 11 post-randomization; **Figure 1A**). Clinician-collected vaginal swabs were obtained at the pre-MTZ and post-MTZ visits prior to randomization, and at weeks 4, 8, 12, and 24 post-randomization. Detailed demographic, clinical, and behavioral data showed no notable differences between arms (Cohen et al., 2020). The trial enrolled and randomized 228 participants (152 in the LBP arm and 76 in the placebo arm); samples from 213 of these participants (142 in the LBP arm and 71 in the placebo arm) were available for this analysis, representing a total of 1,156 unique study visits.

**Figure 1:**
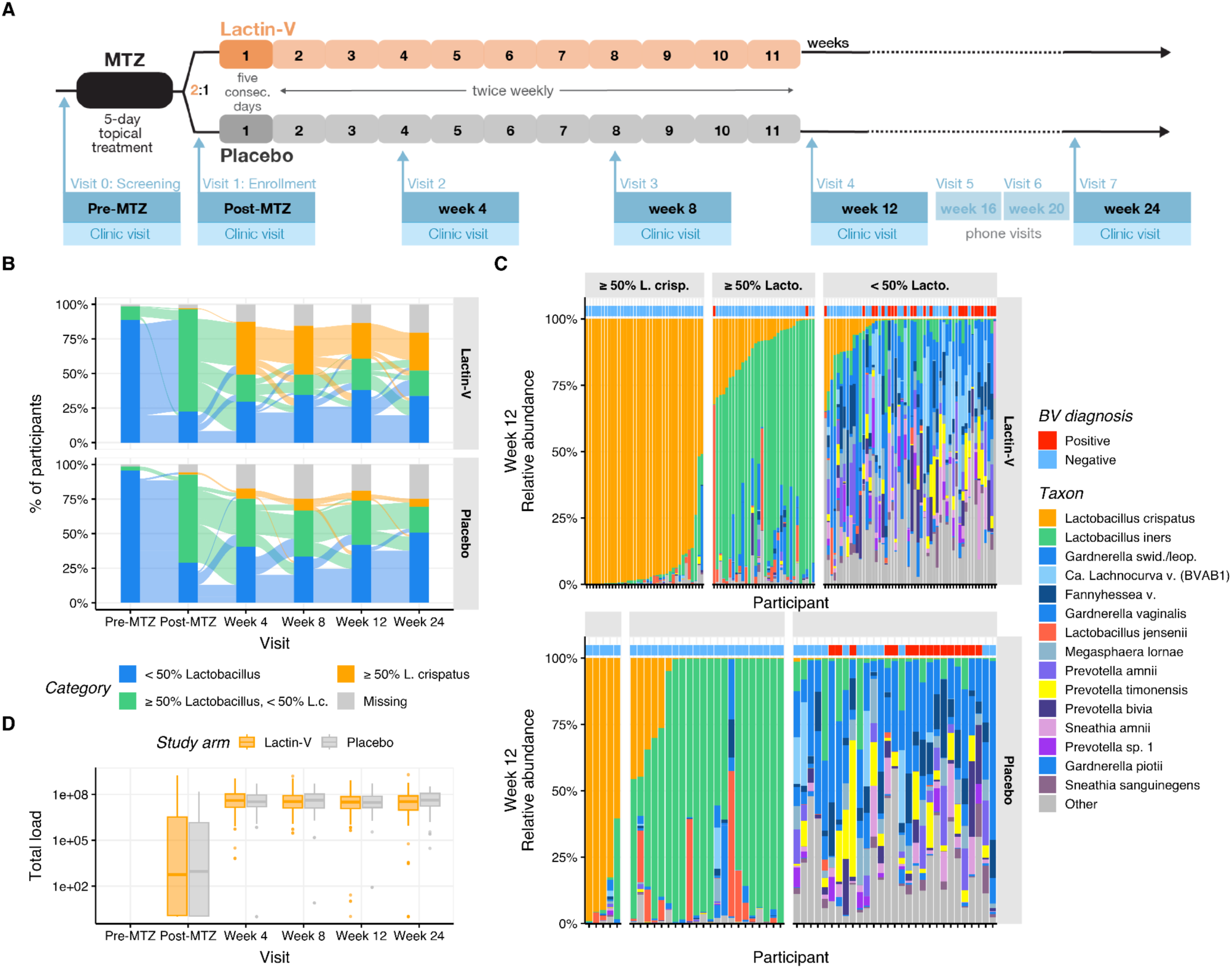
LACTIN-V trial design and microbiota endpoints. A. Schematic of LACTIN-V trial study design and sampling visit timing. B. Participants were grouped into three microbiota categories: *L. crispatus*-dominant colonization (≥ 50% *L. crispatus* relative abundance), *Lactobacillus* (non-*crispatus*)-dominant colonization (≥ 50% *Lactobacillus,* < 50% *L. crispatus*), or non-*Lactobacillus*-dominant (< 50% *Lactobacillus*) based on results of bacterial 16S rRNA gene sequencing. The Sankey diagram displays how participants transitioned between these three categories throughout the trial. C. Microbiota composition and BV diagnosis at week 12 for each participant in the LBP and placebo treatment arms. Relative abundances of the 15 most abundant taxa are shown, with remaining taxa summed as “Other”. Participants are grouped by treatment arm into the categories defined as in Figure 1B, with BV diagnosis indicated along the top margin. D. Total bacterial load (measured by qPCR) at each scheduled visit in each arm. Boxplots (here and in subsequent figures) represent the median (middle horizontal line), the 25th and 75th percentiles (lower and upper boundaries of boxes, respectively, “IQR”), measurements that fall within 1.5 times the IQR (whiskers), and any individual measurement outside that range (dots).

### LBP Effects on *L. crispatus* and total *Lactobacillus* colonization

Bacterial microbiota composition was determined by bacterial 16S V4 ribosomal RNA (rRNA) gene sequencing. Four samples were excluded due to technical failures of PCR amplification and sequencing (<10^3^ processed reads per sample). Of the remaining 1,152 samples (**Table S1, Figure S1A**), the median number of analyzable reads per sample was 31,177 (IQR 23,774-41,251). No significant differences in microbiota composition were observed between study arms at either the pre-MTZ or post-MTZ visits (PERMANOVA *p* = 0.63 and *p* = 0.14, respectively; **Figure S1B-C, SI**).

To assess microbiota impacts of LBP treatment, we defined two outcome parameters based on microbiota proportional composition: ≥50% relative abundance of *L. crispatus* (“*L. crispatus-*dominant”) and ≥50% summed relative abundance of *Lactobacillus* genus (“*Lactobacillus*-dominant”, including *L. crispatus*). The primary microbiota endpoint for this analysis was establishment of an *L. crispatus*-dominant vaginal community at week 12, the timepoint corresponding to the trial’s primary clinical endpoint (Cohen et al., 2020). Secondary microbiota endpoints were *L. crispatus*-dominance at week 24 and total *Lactobacillus*-dominance at weeks 12 or 24. Almost all participants in both treatment and placebo arms had non-*Lactobacillus*-dominant microbiota (<50% *Lactobacillus*) at the pre-MTZ screening visit, consistent with the fact that all had met criteria for clinical BV (**Figure 1B**). The majority in both arms transitioned to *Lactobacillus*-dominance at the post-MTZ visit that was largely driven by high relative abundance of *L. iners* (**Table S2**). Only one participant in each arm exhibited *L. crispatus*-dominant colonization at the post-MTZ visit (**Figure 1B**). Microbiota composition of the two arms diverged after randomization, with 30% of LBP recipients (n=37) compared to only 9% of placebo recipients (n=5) achieving *L. crispatus*-dominant colonization at week 12 (the primary endpoint), resulting in a benefit ratio of 3.4 (95% CI: 1.4 - 8.1; p < 0.005; **Table 1**, **Figure 1B-C**). At week 24, 35% of LBP recipients (n=39) versus just 8% of placebo recipients (n=4) achieved *L. crispatus*-dominant colonization, for a benefit ratio of 4.5 (95% CI: 1.7 - 11.9). The LBP did not increase rates of total *Lactobacillus*-dominant colonization at week 12 compared to placebo (56% and 48% of LBP and placebo recipients, respectively; benefit ratio: 1.16; 95% CI: 0.85 - 1.59), but did at week 24 (58% and 33% of LBP and placebo recipients, respectively; benefit ratio: 1.76; 95% CI: 1.15 - 2.68; **Table 1**, **Figure 1B-C**).

**Table 1:** Benefit ratios for LBP microbiota effects.

| Taxon | Week | Microbiota Endpoint | LACTIN-V | Placebo | Benefit Ratio (95% CI) | P Value |
| --- | --- | --- | --- | --- | --- | --- |
| *L. crispatus* | Week 12 | ≥ 50% *L.c.* | 37 (30%) | 5 (9%) | 3.37<br>(1.40 - 8.11) | < 0.005 |
|  |  | < 50% *L.c.* | 86 (70%) | 51 (91%) |  |  |
|  | Week 24 | ≥ 50% *L.c.* | 39 (35%) | 4 (8%) | 4.49<br>(1.69 - 11.90) |  |
|  |  | < 50% *L.c.* | 74 (65%) | 48 (92%) |  |  |
| *Lactobacillus* | Week 12 | ≥ 50% *Lacto* | 69 (56%) | 27 (48%) | 1.16<br>(0.85 - 1.59) |  |
|  |  | < 50% *Lacto* | 54 (44%) | 29 (52%) |  |  |
|  | Week 24 | ≥ 50% *Lacto* | 65 (58%) | 17 (33%) | 1.76<br>(1.15 - 2.68) |  |
|  |  | < 50% *Lacto* | 48 (42%) | 35 (67%) |  |  |

Benefit Ratios and associated 95% confidence intervals were computed using Wald’s method.

We next investigated how these microbiota parameters related to the clinical diagnosis of BV by comparing clinical and microbiota results for all post-randomization visits (weeks 4 and beyond, n = 729 visits). Among the 209 visits in which participants had ≥50% *L. crispatus* colonization, none had concurrent clinical BV (**Table S3** and **Figure 1C**), and among 194 visits with <50% *L. crispatus* but ≥50% total *Lactobacillus*, only 1% (n=2) had BV. However, of the 326 visits with <50% total *Lactobacillus*, 50% (n=164) had clinical BV (**Table S3**). Thus, *Lactobacillus*-dominance (including *L. crispatus*-dominance) was highly specific for the absence of BV, while non-*Lactobacillus*-dominance was highly sensitive but less specific for BV.

In addition to sequencing-based microbiota composition analysis, total bacterial load in vaginal swab samples was assessed via quantitative PCR (qPCR) at the post-MTZ and post-randomization visits. Median bacterial load was significantly lower and more variable at the post-MTZ visit (median 10^2.69^ copies/swab; IQR 10^0.08^ - 10^6.44^) than at subsequent visits (median 10^7.52^ copies/swab; IQR 10^7.09^ - 10^7.93^ for all subsequent visits), consistent with antibiotic-mediated depletion of vaginal microbiota biomass during BV treatment, followed by bacterial repopulation of the vaginal environment (**Figure 1D**). We assessed whether post-MTZ microbiota composition (as determined by 16S rRNA gene sequencing) was correlated with total bacterial load using a multi-table multivariate generalization of the squared Pearson correlation, the RV coefficient (Josse and Holmes, 2016; Robert and Escoufier, 1976). This analysis revealed no significant correlation between post-MTZ microbiota composition and bacterial load (RV coefficient = 0.02, *p*-value > 0.1).

### Microbiota composition trajectories throughout the trial

To characterize microbiota trajectories with greater resolution, we summarized microbiota composition using topic mixtures (Sankaran and Holmes, 2019; Symul et al., 2023). Compared to clustering or categorizing samples into community state types (CSTs) (Ravel et al., 2011), cervicotypes (CTs) (Gosmann et al., 2017), or sub-CSTs (France et al., 2020), this method provides probabilistic (rather than binary) weightings, allowing for more accurate descriptions of sample taxonomic composition and improved characterization of longitudinal dynamics (Symul et al., 2023). In our cohort, assignment to reference CST performed poorly (median Bray-Curtis dissimilarity with assigned CSTs > 0.33, **SI**), especially for *Prevotella*-dominated samples (median BC dissimilarity > 0.5, **SI**), and with over 40% of samples being equally similar to two or more CSTs (**SI**). To identify topics, which can be interpreted as bacterial subcommunities (*i.e.,* consortia of bacterial taxa that co-occur with each other or with other taxa), and estimate their proportions in each sample, we fitted a latent Dirichlet allocation (LDA) model (a Bayesian “topic model”) (Blei et al., 2003) to the taxonomic counts. We adapted a previously described approach (Symul et al., 2023) in which topics were constrained to be fully composed of either *Lactobacillus* or non-*Lactobacillus* species. Four non-*Lactobacillus* topics were inferred from the data (**Figure S2A, B**) and we defined four *Lactobacillus*-dominated topics, including three exclusively composed of a single *Lactobacillus* species (*L. crispatus*, *L. iners*, and *L. jensenii*/*mulieris*) and one composed of a mixture of the remaining *Lactobacillus* species (**Figure S2C**). **Figure 2A** shows inferred composition of each non-*Lactobacillus* topic, while **Figure 2B** displays topic proportions for each participant throughout the trial.

**Figure 2:**
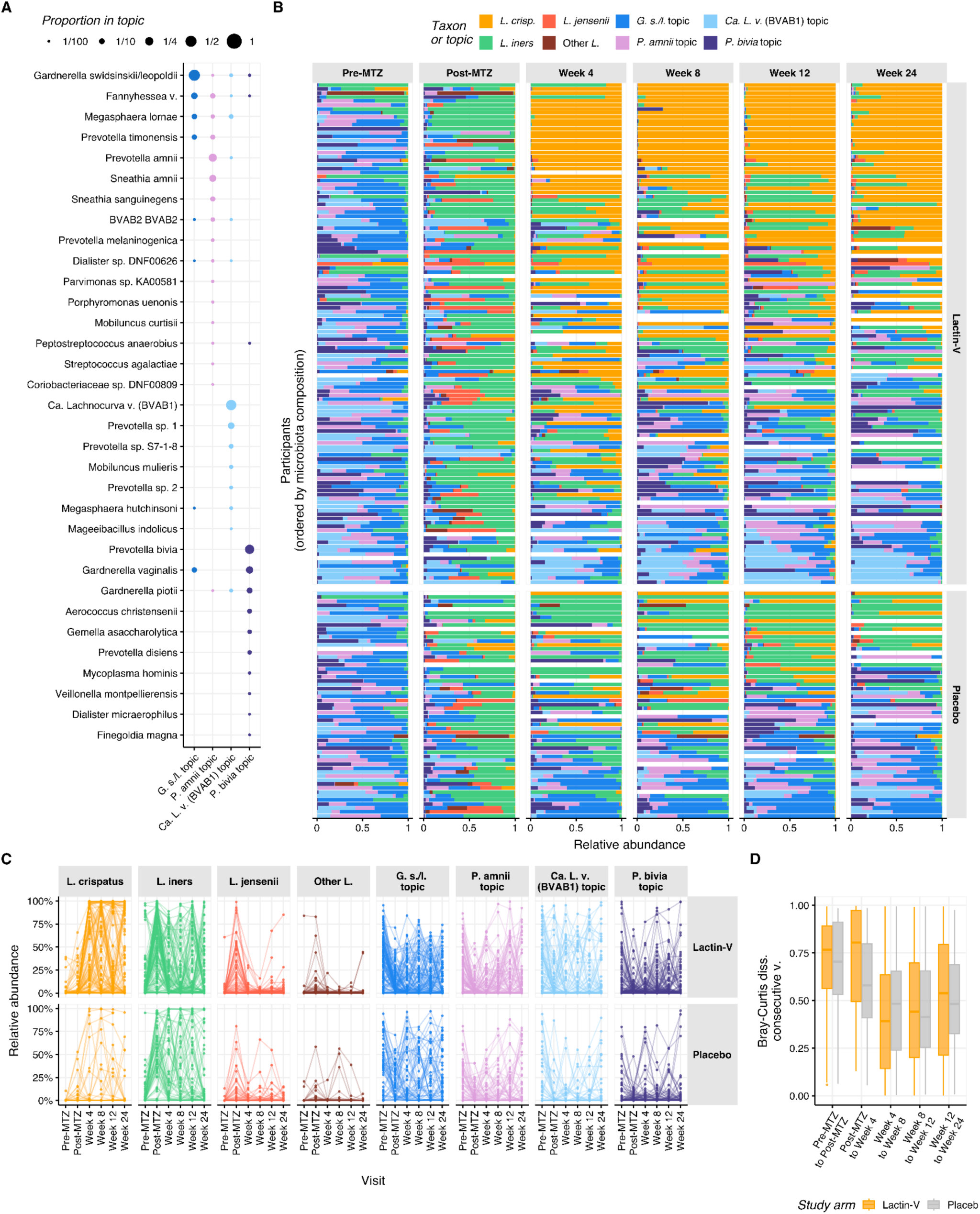
Microbiota trajectories throughout the trial. A. Proportion of each taxon (y-axis) in each non-*Lactobacillus* topic (x-axis) as estimated by Latent Dirichlet Allocation (LDA) from 16S rRNA gene sequencing data. Proportions sum to one for each topic. Taxa were included if they made up at least 1% of any topic and non-*Lactobacillus* topics are named according to their most predominant taxon. “*G. s/l*”: *Gardnerella swidsinskii-leopoldii*. “*P. amnii*”: *Prevotella amnii*. “*Ca.* L. v. (BVAB1)”: *Candidatus* Lachnocurva vaginae (BVAB1). “*P. bivia*”: *Prevotella bivia*. B. Microbiota composition expressed as topic relative abundance (x-axis) for each participant at each scheduled visit. Results are depicted for participants with microbiota compositional data for at least three of the six visits. Results are grouped by visit (horizontal panels) and treatment arm (vertical panels), and the y-axis is ordered by microbiota composition trajectories (see **SI**). C. Relative abundance of each topic (horizontal panels) in each arm (vertical panels) at each visit. Samples from the same participant are connected by a line. D. Bray-Curtis dissimilarity between each participant’s microbiota composition at the two indicated consecutive visits, calculated from 16S rRNA gene amplicon sequence variant (ASV) relative abundances. For each pair of consecutive visits, values are shown only for participants with available microbiota data from both visits.

Three non-*Lactobacillus* topics were highly prevalent and abundant in both arms at the pre-MTZ visit, including topics in which the predominant taxa were *Candidatus* Lachnocurva vaginae (BVAB1), *Gardnerella swidsinskii/leopoldii*, and *Prevotella amnii* (**Figure 2C** and **S2D**). Microbiota composition shifted substantially at the post-MTZ visit (**Figure 2D**), where besides high *L. iners* relative abundance, the non-*Lactobacillus* topic with the highest relative abundance was the topic in which *Gardnerella swindsinskii/leopoldii* was predominant (**Figure 2C** and **S2D**). Another major shift in microbiota composition occurred between the post-MTZ visit and the week 4 visit, which was larger in the LBP arm (median pairwise Bray-Curtis dissimilarity: 0.81; IQR: 0.50 - 0.97) than the placebo arm (median Bray-Curtis dissimilarity: 0.58; IQR 0.41 - 0.80), driven primarily by increased *L. crispatus* abundance among LBP recipients (**Figure 2C-D** and **S2D**). Microbiota composition was more stable after week 4, with median Bray-Curtis dissimilarity <0.55 at week 8 and all subsequent visits, which was comparable between arms (**Figure 2D**). LBP recipients who achieved *L. crispatus*-dominant colonization had more stable microbiota than their counterparts (**Figure S2E, SI**).

The observation that microbiota composition became more stable after week 4 was also supported by longitudinal patterns in microbiota categories. Among 115 participants with available data from both week 4 and week 12 visits, 71% (25 of 35; 95% CI: 53%-85%) of LBP recipients who achieved ≥50% *L. crispatus* at week 12 also had *L. crispatus*-dominance at week 4 (**Figure 2B, Table S4**). Similarly, 71% (27 of 38; 95% CI: 54%-84%) of LBP recipients who achieved *L. crispatus*-dominance at week 24 also had *L. crispatus*-dominance at week 4 (**Figure 2B**, **Table S5**). However, early *L. crispatus*-dominant colonization did not guarantee persistence, as only 49% (25 of 51) and 57% (27 of 47) of LBP recipients with *L. crispatus*-dominance at week 4 retained it at weeks 12 and 24, respectively. By contrast, among 38 LBP recipients with non-*Lactobacillus*-dominant microbiota at week 4, 79% remained non-*Lactobacillus* dominant at week 12 compared to just 10.5% each with *L. crispatus*-dominance or other *Lactobacillus*-dominant colonization (**Table S4**). Thus, microbiota communities established early tended to persist and early *Lactobacillus*-dominance – particularly *L. crispatus*-dominance – was strongly linked to ongoing *Lactobacillus*-dominance.

### Metagenomic identification of the CTV-05 strain and assessment of *L. crispatus* strain dynamics

The observation that LBP treatment promoted *L. crispatus* colonization prompted us to examine what fraction of *L. crispatus* observed in LBP recipients represented the CTV-05 strain versus non-LBP native strains. We used metagenomic sequencing to assess *L. crispatus* strain dynamics, with strain genotypes and proportions inferred using StrainFacts (Smith et al., 2022). In a separate analysis, simulations and experimental spike-in data showed that StrainFacts performs well at inferring LBP (including CTV-05) strain genotypes and proportions in samples with ≥5% overall relative abundance of *L. crispatus* (Shih et al., 2025). In the LACTIN-V trial samples, *L. crispatus* strain inference was successful in 313 samples, identifying 24 genotypically distinct strains with at least 10% fractional strain abundance in at least one sample. To determine which strain represented CTV-05, we generated a closed assembly of the CTV-05 genome, which nearly perfectly matched the StrainFacts genotype of a widely prevalent inferred strain within the metagenomic dataset (Jaccard similarity >0.999). Among 57 samples with inferred *L. crispatus* strain composition from placebo recipients (any visit) or from LBP recipients prior to LBP administration, StrainFacts inferred CTV-05 presence in just 5 total samples from 5 unique participants (**Figure 3A**). This result suggested our strain inference approach had a low false positive CTV-05 detection rate; we cannot rule out that some “false positives” represented native strains with high genotypic similarity to CTV-05. Collectively, these results support that CTV-05 detection was achieved with high accuracy in sample metagenomes.

**Figure 3:**
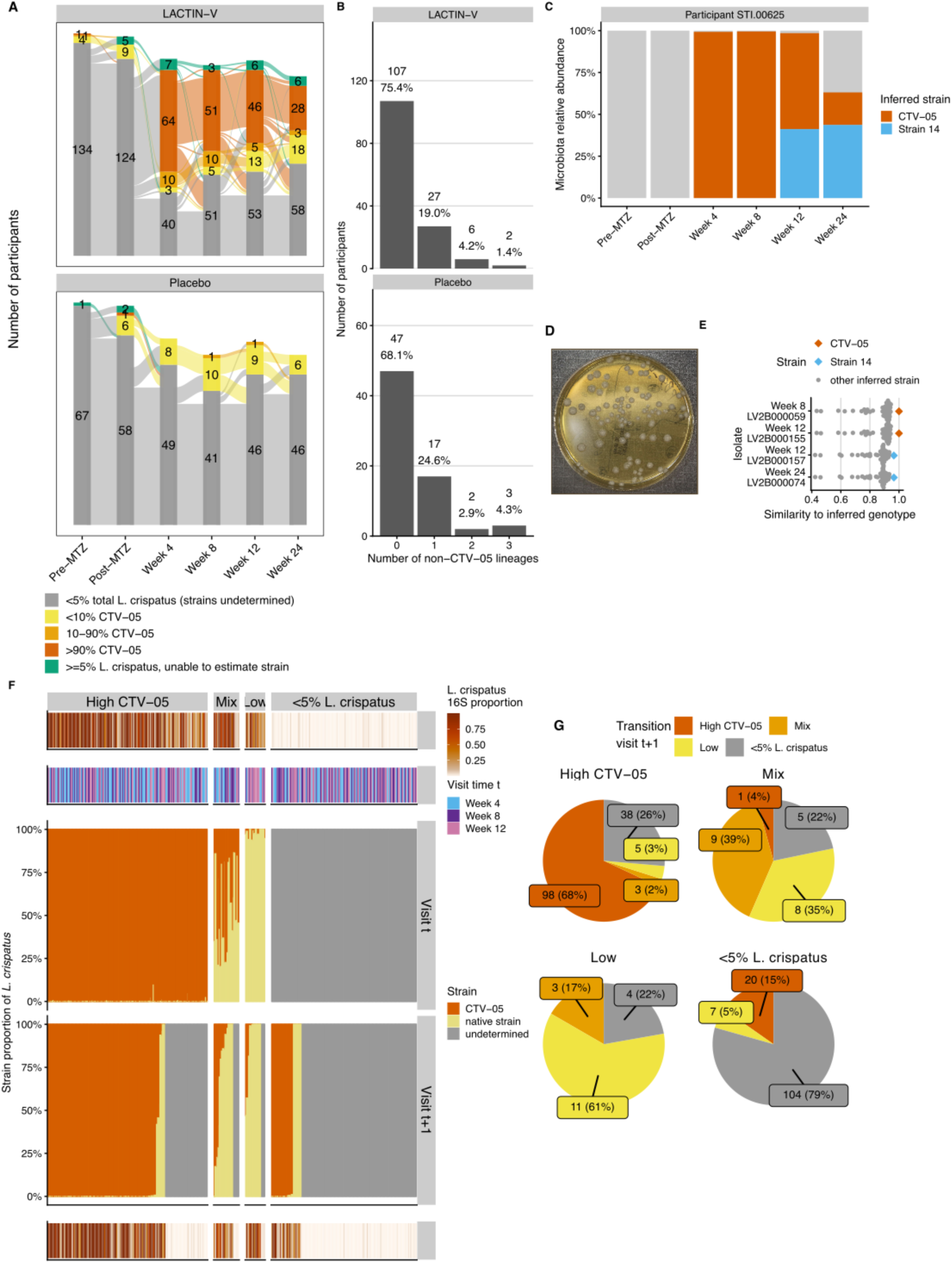
*L. crispatus* strain dynamics among LBP recipients. A. Sankey diagram showing dynamics of CTV-05 versus non-CTV-05 *L. crispatus* strain colonization among LBP recipients. Strains were inferred from shotgun metagenomic whole genome sequencing (mWGS) using StrainFacts for all samples with ≥5% total *L. crispatus* relative abundance as determined from 16S rRNA gene sequencing. Samples were categorized based on proportional *L. crispatus* strain abundance as >90% CTV-05 (“High CTV-05”), 10-90% CTV-05 (“Mixed”), <10% CTV-05 (“Low CTV-05”). Strain inference was unsuccessful for a small fraction of samples (depicted in green) due to technical failures of metagenomic sequencing. B. Histograms showing the number (percentage) of study participants in each treatment arm with the indicated number of inferred native (non-CTV-05) *L. crispatus* strains detected at least once during the trial. C. Metagenomically inferred *L. crispatus* strain composition throughout the trial in an example participant from whom targeted isolations were performed at weeks 8, 12, and 24. The plot depicts overall microbiota relative abundance, with non-*L. crispatus* species summed (gray) and inferred *L. crispatus* strains shown individually in red (CTV-05 strain) and blue (a native strain: “Strain 14”). D. Example *Lactobacillus* MRS agar plate from bacterial isolations showing characteristic *L. crispatus* colony morphology. E. Genomes of *L. crispatus* isolated from the week 8, 12, and 24 visits of the participant in Figure 3C identified two strains nearly identical to the metagenomically inferred strains in these samples (assessed by Jaccard similarity to StrainFacts-inferred genotypes) and dissimilar from all other inferred strains in the cohort, none of which were detected in this participant’s samples. The isolated strains included the CTV-05 strain isolated from week 8 and 12 samples (red), and native Strain 14 isolated from week 12 and 24 samples (blue; see also **Figure S3A**). F. Transition plot for LBP recipients showing total *L. crispatus* relative abundance (determined by 16S rRNA gene sequencing, top and bottom rows) and strain category proportions (third and fourth rows) at each pair of consecutively scheduled post-randomization visits with available strain inference data or at which strain inference was not performed due to <5% total *L. crispatus* relative abundance. Strain categories in the third and fourth rows show proportions of observed *L. crispatus* representing CTV-05 versus the sum of the native strain(s) at each visit; “undetermined” refers to samples in which strain inference was not performed. Pairs including a visit in which strain inference was unsuccessful due to technical failures of metagenomic sequencing are not shown. For each pair of visits, the top three panels show *L. crispatus* relative abundance, visit week, and strain category at the first visit in the pair (“Visit t”) and the bottom two panels show strain category and relative abundance at the subsequent visit (“Visit t+1”). Visit pairs are grouped along the horizontal axis by strain category at Visit t and ordered within groups by CTV-05 strain proportion at Visit t+1. G. Pie charts for each Visit t strain category from Figure 3F, summarizing subsequent *L. crispatus* strain category frequencies at Visit t+1.

To further characterize fidelity of the strain-tracing approach, we analyzed inference results for native *L. crispatus* strains, supplemented by targeted bacterial isolations and genome sequencing to experimentally test bioinformatic predictions. Most participants in whom a native strain was inferred exhibited colonization by only a single native strain at any point during the study, with a few participants colonized by two or three native strains (**Figure 3B**). Since StrainFacts inferences are performed blinded to whether samples are derived from the same participant (Shih et al., 2025; Smith et al., 2022), consistent detection of the same 1-3 native strains in multiple samples from the same participant additionally supported validity of the inferences. We also performed targeted bacterial isolations of *L. crispatus* from a subset of samples inferred to contain CTV-05, one or more native strains, or a mixture of CTV-05 and native strains. The resulting isolates were highly genotypically similar to inferred strains from the same samples, including both CTV-05 and native strains (results for an example participant shown in **Figure 3C-E**, with additional participants detailed in **Figure S3A**). These experimental results further validated our metagenomic strain inference approach.

We examined *L. crispatus* strain dynamics by classifying samples with inferred strain data into three categories: samples with >90% CTV-05 fractional strain abundance (“High-CTV-05”), samples with 10-90% CTV-05 (“Mixed”), and samples with <10% CTV-05 (“Low-CTV-05”; **Figure 3A**). These thresholds were defined based on simulations to determine parameters maximizing reliability of strain detection (Shih et al., 2025). At each post-randomization timepoint, the majority of LBP recipients with analyzable *L. crispatus* strains had high-CTV-05 strain abundance, but the proportion with high-CTV-05 or mixed strains decreased over time, while the proportion with high native *L. crispatus* strain abundance (*i.e.*, low-CTV-05) increased (**Figure 3A**). At week 4, 96.1% (n=74) of LBP recipients with ≥5% *L. crispatus* had high-CTV-05 or mixed strains, while just 3.9% (n=3) had high native strains. However, by week 24 only 63.2% (n=31) had high-CTV-05 or mixed strains while those with high native strains increased to 36.8% (n=18). To characterize this phenomenon in greater detail, we examined CTV-05 fractional strain abundance for each pair of consecutive post-randomization visits (**Figure 3F-G**). Among 144 visits with high-CTV-05, two main outcomes were observed at the next visit: 68% (n=98) maintained high-CTV-05 and 26% (n=38) transitioned to <5% total *L. crispatus* colonization (strain composition undetermined). Among the 18 visits with high native strain abundance, 61% (n=11) retained high native strain abundance at the next visit and none transitioned to CTV-05 dominance. Interestingly, among 23 visits with mixed CTV-05/native strain composition, only one (4%) transitioned to high-CTV-05, while 39% (n=9) remained mixed and 35% (n=8) transitioned to high native strain abundance, including some transitions to high native strain abundance that occurred while participants were still receiving LACTIN-V (**Figure S3B**). These results show CTV-05 was frequently the sole or dominant *L. crispatus* strain in LBP recipients, but when native strains dominated or co-occurred with CTV-05, they often replaced CTV-05 at subsequent visits but were almost never replaced by it.

### Intervention effects on vaginal inflammation

We assessed the effects of MTZ and LBP treatment on vaginal mucosal immune state and inflammation by measuring cytokine and chemokine concentrations in vaginal swab eluates using a previously described custom Luminex assay (Gosmann et al., 2017; Symul et al., 2023). Concentrations of 18 cytokines and chemokines were log-transformed for variance stabilization and imputed when outside their respective limits of quantification (**Figure S4A**). Concentrations of all chemokines and cytokines were strongly correlated, with the first principal component (PC1) accounting for 50% of the total variation (**Figure S4B**). This phenomenon suggested a size-effect (Jolicoeur and Mosimann, 1960) that we attributed to the (unmeasured) variation in biomass collection between swabs, which was addressed through PC1-subtraction (**SI**). Seven cytokines were excluded from analysis because a large proportion of values were outside the quantification range (**Figure S4A**). Mucosal immune dynamics during treatment were determined by comparing adjusted concentrations of each cytokine or chemokine from each participant’s baseline (pre-MTZ) visit with concentrations at subsequent visits.

We first examined how chemokines and cytokines with known relationships to BV and vaginal microbiota composition (Anahtar et al., 2015; Gosmann et al., 2017; Masson et al., 2019, 2014) changed during treatment. IL-1β concentrations decreased post-MTZ in both arms, while IP-10 showed the opposite pattern, consistent with previously reported pattern with BV treatment (Masson et al., 2014) (**Figure 4A-B**). These effects persisted in both arms through week 12, but IL-1β and IP-10 returned to pre-MTZ levels by week 24 in placebo recipients, leading to a difference of −0.39 (95% CI: −0.66 – −0.14) between adjusted concentrations in the LBP versus placebo arm for IL-1β and of 0.51 (95% CI: 0.21 - 0.79) for IP-10. By contrast, IL-6 concentrations – which have been reported not to differ between women with and without BV (Masson et al., 2014) – remained unchanged from pre-MTZ levels in both treatment arms throughout the study (**Figure 4C**). Sensitivity analyses examining the data prior to adjustment for size-effect via PC1-subtraction revealed findings that were qualitatively similar, but with reduced magnitude (**SI**).

**Figure 4:**
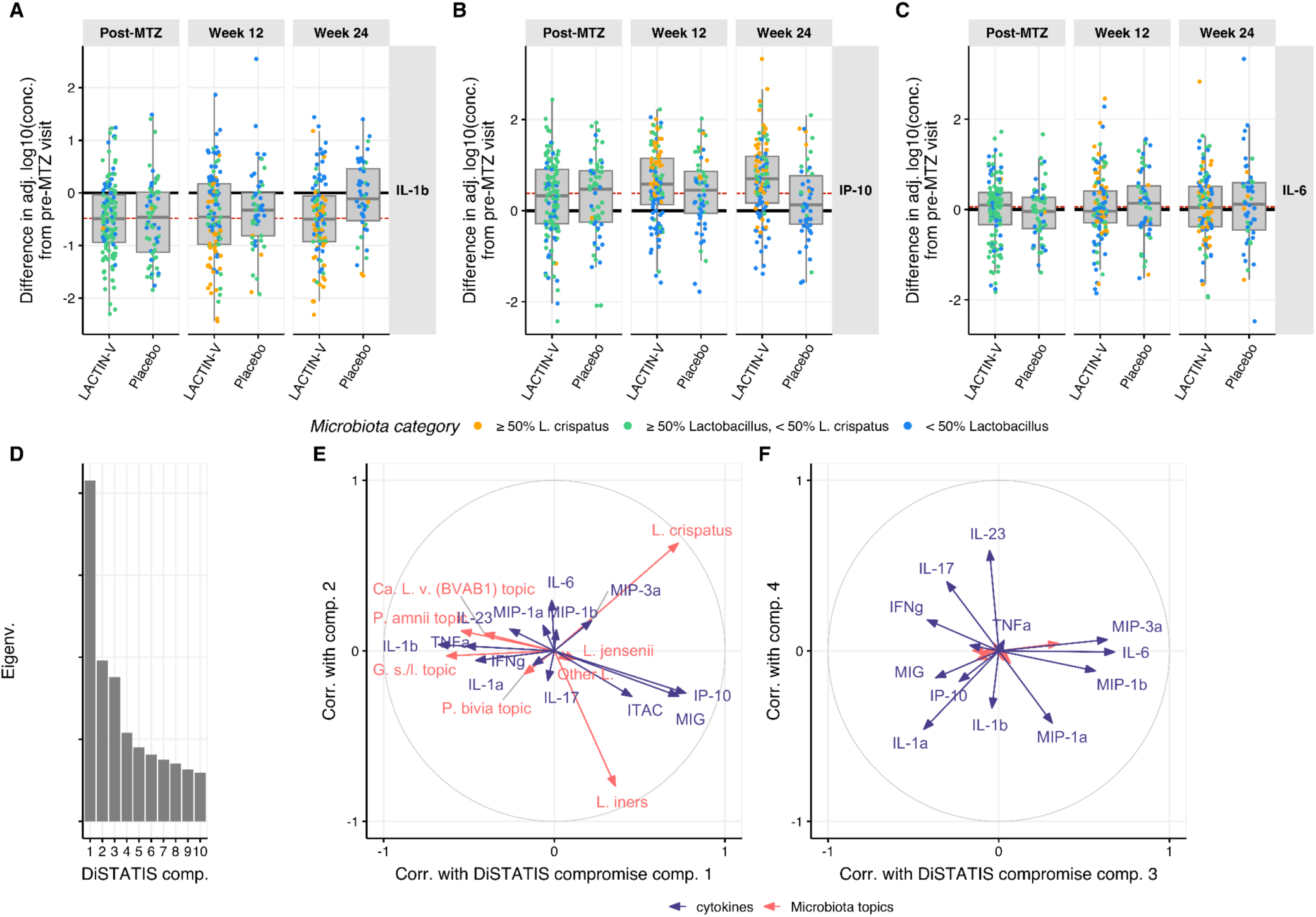
Treatment effects on mucosal inflammation and relationship to microbiota composition. A. Differences in log10-transformed, adjusted IL-1β concentrations between participants’ pre-MTZ visit and their ensuing post-MTZ, week 12, and week 24 visits (horizontal panels) in both treatment arms. Each dot represents a single visit per participant. B. Same as Figure 4A. for IP-10 adjusted concentrations. C. Same as Figure 4A. and **4B** for IL-6 adjusted concentrations. Similar displays are available for all cytokine adjusted concentrations and non-adjusted concentrations in the **SI**. D. DiSTATIS screeplot: eigenvalues (a.u.) of DiSTATIS compromise for the first 10 latent components. E. DiSTATIS correlation circle showing the correlations between the 1st (x-axis) and 2nd (y-axis) DiSTATIS compromise latent dimension and the microbiota topic proportions (red) or cytokine transformed concentrations (blue). F. Same as Figure 4E for DiSTATIS 3rd and 4th latent components.

The reversion of treatment-induced effects for IL-1β and IP-10 concentrations in placebo recipients by week 24 suggested an association with total *Lactobacillus* relative abundance, which differed between treatment arms at this visit (**Table 1**). We therefore examined the correlation of microbiota composition (summarized by topic proportions) with mucosal cytokine/chemokine concentrations. Their RV coefficient was 0.18 with p-value <0.005 (permutation test), indicating significant correlation. Similar RV coefficient values were obtained when repeating analysis independently for each visit (**Figure S4C**) except for a substantially lower correlation at the post-MTZ visit (where microbiota absolute abundance varied widely, **Figure 1D**), suggesting microbiota-immune correlation was not driven primarily by a participant effect. To characterize the microbiota-immune relationship in greater detail, we employed DiSTATIS (Abdi et al., 2009, 2005), a method that uses between-sample dissimilarity matrices to infer a consensus matrix across two datasets and associated latent components. Projecting microbiota topic relative abundances and transformed cytokine/chemokine concentrations onto this latent subspace using correlation circles revealed that the first latent dimension discriminated *Lactobacillus*-dominated samples from non-*Lactobacillus*-dominated samples (**Figure 4D-F**). Chemokines MIG (CXCL9), IP-10 (CXCL10), and ITAC (CXCL11) had high positive correlation along the first latent dimension, indicating positive covariation with *Lactobacillus*, while IL-1β, TNF-α, and IL-1α had high negative correlations, indicating positive covariation with non-*Lactobacillus* species (**Figure 4E**). The second latent dimension discriminated *L. crispatus* from *L. iners* but most cytokines and chemokines had little or no correlation with that component, indicating minimal association of inflammatory markers with individual *Lactobacillus* species (**Figure 4E**). Most cytokines and chemokines had high correlations with the 3rd and/or 4th latent components but the microbiota topics did not, indicating a degree of immune variation that was microbiota-independent (**Figure 4F**). In these dimensions, IFNγ and IL-17 correlated with each other but were anti-correlated with IL-6, MIP-3α, MIP-1β, and MIP-1α. Notably, although IP-10 levels were previously reported to differ in a small subset of LACTIN-V recipients based on whether participants were highly colonized by CTV-05 versus non-CTV-05 *L. crispatus* strains (as measured by qPCR) (Armstrong et al., 2022), we observed no clear differences in microbiota correlation with cytokine/chemokine levels (including IP-10) in analysis that distinguished CTV-05 from other *L. crispatus* strains (**Figure S4D-G**).

### Heterogeneity in LBP treatment effect based on baseline microbiota composition

We next performed exploratory *post-hoc* analyses to investigate whether differences in pre-MTZ microbiota composition corresponded to differences in response to the LBP (*i.e.,* heterogeneity of treatment effect). We stratified participants by their most abundant pre-MTZ taxon (aggregated at the genus-level), identifying four groups for analysis: *Lactobacillus*-predominant, *Gardnerella*-predominant, *Prevotella*-predominant, and *Ca.* Lachnocurva vaginae (BVAB1)-predominant. LBP treatment benefit was evaluated for each group with respect to two distinct outcomes: ≥50% *L. crispatus* colonization (at week 12 or 24) and BV recurrence (by week 12 or 24) (**Figure 5A**). Heterogeneity in benefit among pre-MTZ microbiota groups (**Figure 5B**) was tested using an analysis of deviance comparing nested logistic regression models allowing (or not) for heterogeneity and adjusting for multiple testing by controlling for the false discovery rate (Methods, **SI**).

**Figure 5:**
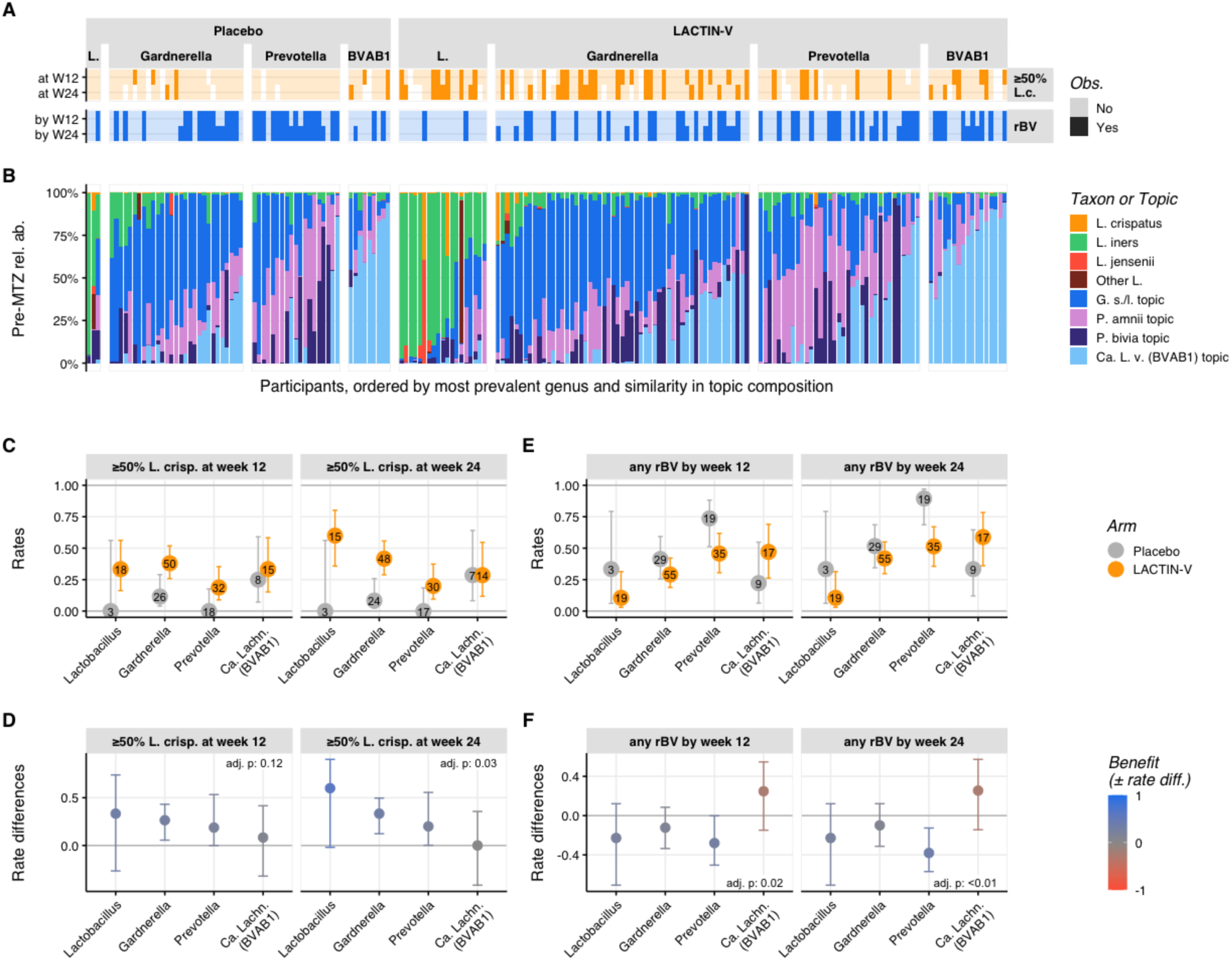
Heterogeneity in intervention effects with respect to pre-MTZ microbiota. A. Participant-level observed outcomes for attainment of *L. crispatus*-dominance at weeks 12 and 24 (dark orange: ≥50% *L. crispatus*; light orange <50% *L. crispatus*) and BV recurrence by weeks 12 and 24 (dark blue: rBV; light blue: no rBV) for each participant with week 12 and/or week 24 data. White indicated data not available. Participants are grouped according to treatment arm and, within treatment arm, by the most predominant bacterial genus at their pre-MTZ visit (“L.” indicates *Lactobacillus*; “BVAB1” indicates *Ca.* Lachnocurva vaginae). Predominant-genus groupings lacking at least two participants in each treatment arm were excluded. B. Pre-MTZ microbiota composition for each participant in Figure 5A (x-axis ordered and vertically aligned as in **5A**), expressed as topic proportions (see Figure 2A for topic definitions). C. Rates and associated 95% CI of *L. crispatus*-dominance (≥50% *L. crispatus*) at week 12 or 24 (horizontal panels) by pre-MTZ microbiota group in each arm. Participants are grouped based on their most predominant genus as defined in Figure 5A-B. CI are computed using Wilson’s scores due to low sample sizes in some groups. D. Rate differences (rate in LBP arm - rate in placebo arm) and associated 95% CI (Wilson’s score method) between arms for results in Figure 5C. Color scale shows degree of benefit (defined as the difference in rates between the two arms) in achieving *L. crispatus*-dominance, with blue indicating benefit with LBP treatment and red indicating benefit with placebo. *P*-values adjusted for multiple testing (across panel **5D** and **5F**) using the Benjamini-Hochberg correction of the analysis of deviance test for heterogeneity in treatment effects when stratifying participants according to their most predominant genus at the pre-MTZ visit are shown in upper right corner. E. Same as (Figure 5C), but for BV recurrence by week 12 or 24. F. Same as (Figure 5D), but for BV recurrence by week 12 or 24. Color scale shows degree of benefit in reducing rBV, with blue indicating benefit with LBP treatment and red indicating benefit with placebo.

Analysis of the relationship between pre-MTZ microbiota groups and establishment of *L. crispatus*-dominant colonization suggested some groups benefitted more from LBP treatment than others (adjusted *p*-values = 0.11 at week 12 and 0.03 at week 24; **Figure 5C, D**). LACTIN-V recipients with pre-MTZ predominance of *Lactobacillus*, *Gardnerella*, or *Prevotella* attained higher rates of *L. crispatus*-dominant microbiota at both weeks 12 and 24 compared to their placebo counterparts, with modestly larger rate differences at week 24 (**Figure 5D**). In contrast, participants with pre-MTZ *Ca.* Lachnocurva vaginae (BVAB1) did not differ in *L. crispatus*-dominant colonization between LBP and placebo arms. Analysis of BV recurrence also showed differences in benefit from LBP treatment based on pre-MTZ microbiota (adjusted *p*-values <0.05 at week 12 and <0.01 at week 2; **Figure 5E-F**). Among LBP recipients with pre-MTZ *Prevotella*-predominance, BV recurred in almost all placebo recipients by week 24, compared to just over half of LBP recipients. By contrast, LBP recipients whose most prevalent taxa pre-MTZ was Ca. *Lachnocurva vaginae* (BVAB1) were unique in exhibiting slightly higher absolute rates of rBV than those who received placebo (**Figure 5E-F**). We used similar methods to perform a sensitivity analysis in which participants were grouped based on pre-MTZ CSTs (France et al., 2020) or using a model-based approach relying on topic relative abundances. Findings from this analysis were largely consistent with results of our predominant-taxon analysis above (**Figure S5, SI**). Together, these exploratory analyses showed that benefits of LBP treatment in achieving *L. crispatus*-dominance and preventing BV recurrence differed depending on pre-MTZ microbiota composition, with participants who had pre-MTZ predominance of *Ca.* Lachnocurva vaginae (BVAB1) exhibiting a lack of benefit compared to placebo, whereas other participants experienced more favorable responses.

### Factors associated with achieving *L. crispatus* dominance in LBP recipients

To identify factors associated with establishing *L. crispatus* dominance among LBP recipients, we performed a series of multiblock partial least square discriminant analyses (MB-PLS-DA) on the longitudinal LACTIN-V arm data (**Figure 6A-C**, **S6**, **S7A**). Based on the observation that early *L. crispatus*-dominant colonization correlated with *L. crispatus* colonization at later visits (**Table S4**, **S5**), we analyzed factors from prior visits that associated with the participants’ microbiota category by week 4, as well as separate analyses of factors associated with microbiota category at ensuing visits. We defined three models corresponding to the different phases of the trial: the “initial phase” spanned from the post-MTZ visit to week 4, the “continuation phase” spanned from week 4 to week 12, and the “follow-up phase” spanned from week 12 to week 24 (**Figure S6A**). For each phase, factors of interest (*i.e.,* explanatory variables) were grouped into 13 thematic blocks (**Figure 6A**, **Table S6**). The first block characterized participant demographics, blocks 2-4 described participant baseline vaginal ecosystem (*i.e.*, pre-MTZ microbiota composition and diversity, pH, cytokines concentrations), blocks 5-8 quantified the vaginal ecosystem at the previous visit (*e.g.,* at the post-MTZ visit in the initial phase, when the response is week 4 colonization status), while blocks 9-13 characterized participant sexual behavior, douching/bleeding, antibiotic use, and product adherence (**Table S6**). For each phase, MB-PLS-DA was used to evaluate the ability of these variables to discriminate between the following microbiota categories at each visit starting from week 4: ≥50% *L. crispatus*, ≥ 50% *Lactobacillus* but <50 % *L. crispatus*, or < 50% *Lactobacillus* (see also **Figure 1B**). We also used nested models to assess the additional contribution of specific blocks in discriminating colonization status (**SI**, **Figure S6A-C**).

**Figure 6:**
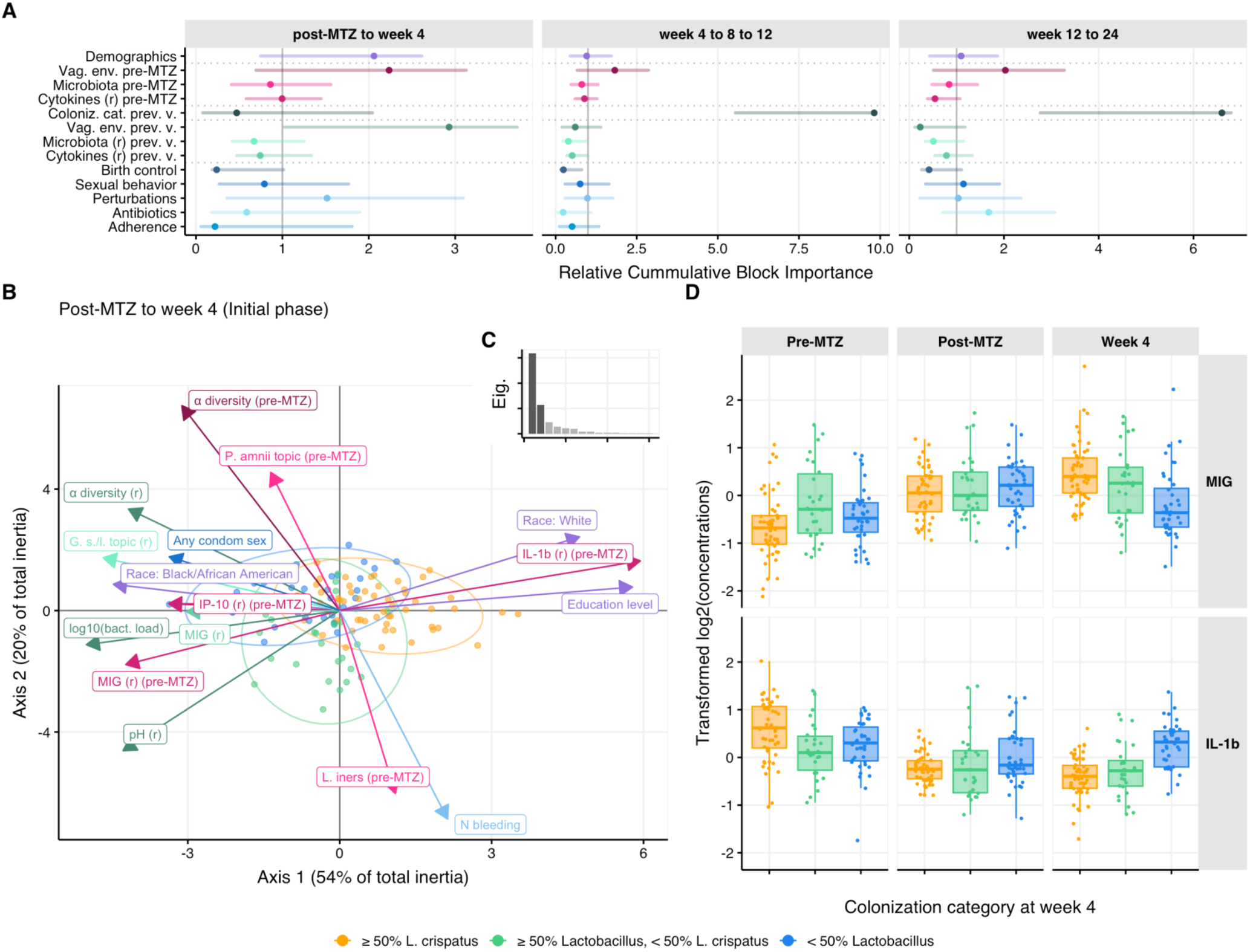
Multiblock analysis of factors associated with microbiota categories in LACTIN-V recipients. A. Multiblock partial least squared discriminant analysis (MB-PLS-DA, **Figure S6A**) of factors predictive of microbiota categories (≥50% *L. crispatus*; ≥50% *Lactobacillus* and <50% *L. crispatus*; or <50% *Lactobacillus*) at the indicated visits. Relative cumulative importance indices (x-axis, Methods) are shown for each thematic block of explanatory variables (y-axis, color) for the initial phase model (post-MTZ to Week 4; left panel), continuation phase model (Week 4 to 8 to 12; middle panel), and follow-up phase model (Week 12 to 24; right panel). Dots indicate point estimates and translucent horizontal bars represent bootstrapped 95% CI. Vertical lines at x = 1 show the expected value under the (null) hypothesis that all variables have the same importance. Block definitions along with the description of the variables included in each block are provided in **Table S6**. Horizontal dashed lines separate blocks included in the nested models (**Figure S6A**, **SI**, Methods). B. Bi-plot for the MB-PLS-DA initial phase model (from post-MTZ to week 4). Dots represent scores (each dot is a participant) while arrows represent loadings (each arrow is an explanatory variable, see **Figure S7A**) for the most important variables (Methods) in the space of the 2 first latent variables. Dots (participants) are colored by their corresponding response categories (*i.e.,* their colonization status at week 4). Perfect separation of the categories would indicate perfect prediction of colonization status at the week 4 visit. Arrows (explanatory variables) are colored by the block to which they belong to - Figure 6A and **Table S6** serve as color legend. C. Scree-plot of the first 15 eigenvalues of the model from Figure 6B. The first two latent variables (selected by cross-validation) are shown in dark gray. D. Transformed log_2_-concentrations (y-axis) of two cytokines (MIG and IL-1b) whose pre-MTZ levels (left panel) were identified as associated with participants’ colonization status at week 4 (x-axis, colors) by the model (Figure 6A and **S7A**). Concentrations at the post-MTZ and week 4 visits are shown in the middle and right panels. Each dot is a participant, colored by the colonization status at week 4 for each panel.

In the model for the LBP arm initial phase, the most important block for discriminating week 4 colonization categories was the previous (*i.e.,* post-MTZ) vaginal environment (**Figure 6A**, left panel). This block included post-MTZ total bacterial load, α-diversity, and pH. Lower values of these three factors were associated with higher chances of ≥50% *L. crispatus* at week 4, and were among the 5 most important variables in the model (**Figure 6B-C** and **S7A**). The next most important blocks were demographics and blocks characterizing participant pre-MTZ vaginal environment, including pre-MTZ vaginal pH, α-diversity, cytokine concentrations, and microbiota composition (as described by topic proportions, **Figure 6A** and **S7A**). Specifically, high pre-MTZ α-diversity and pH were negatively associated with *L. crispatus* colonization at week 4 (**Figure 6B**). Interestingly, pre-MTZ adjusted concentrations of the chemokine IL-1β were positively associated with achieving *L. crispatus*-dominance at week 4 while pre-MTZ IP-10 and MIG levels showed an opposite association (**Figure 6B**, **6D**). Self-declared race and education were the most important demographic block variables, with white and more highly educated participants having higher rates of *L. crispatus*-dominance (**Figure S7A**). However, race and education were highly correlated in the cohort (**Figure S7B**) such that their individual contributions to explaining *L. crispatus* colonization could not be clearly distinguished. While these two demographic variables were also correlated with several pre-MTZ vaginal characteristics that also explained colonization such as pre-MTZ α-diversity (**Figure S7C**), they still mildly contributed to model explanatory power (**Figure S6C**). The remaining blocks or variables were not important for discriminating between initial phase colonization categories (**Figure 6A-B**, **Figure S7A**), possibly because variables in these blocks contributed limited information as overall adherence was high, use of antibiotics and douching was relatively rare, and some birth control options were rare in this cohort (**SI**). In contrast, within these blocks, factors with higher heterogeneity across participants such as bleeding and sexual activity showed greater variable importance (**Figure S7A**).

In the continuation phase model for the LBP arm (from week 4 to week 12), the colonization category at the previous visit was by far the most important block associated with colonization category at the following visit (**Figure 6A**), consistent with higher stability of microbiota composition from week 4 to 12 (**Figure 2D**). While no other blocks significantly improved model performance (**Figure S6C**), individual variables including sexual activity, bleeding, douching, and non-hormonal IUDs were all negatively associated with high-level *L. crispatus* colonization (**Figure S7A**). In the model for the LBP follow-up phase (from week 12 to week 24), the colonization category at the previous visit (*i.e.,* week 12) was again strongly associated with the colonization category at week 24. However, in contrast to the previous phase, sexual behavior and antibiotic use only mildly contributed in explaining week 24 colonization categories (**Figure 6A**, right panel, **Figure S6C**, **S7A**).

Analogous analyses for placebo recipients did not show significant predictive value for either the initial phase or the follow-up phase in cross-validation (**Figure S8A-C**, **S9**). The placebo continuation phase model had modest predictive value, with microbiota category at the previous visit serving as the best predictor of microbiota category at the next visit (**Figure S8A-C**, **S9**).

## Discussion

Treatment with vaginal live biotherapeutic products (LBPs) offers significant promise to improve health, but mechanisms, correlates, and strain dynamics of LBP colonization and efficacy remain incompletely understood (Bradshaw et al., 2025). We performed comprehensive analyses of samples and data from a randomized, double-blind, placebo-controlled phase 2b trial of LACTIN-V, a single-strain *L. crispatus* LBP for prevention of BV recurrence (Cohen et al., 2020). We employed microbiome sequencing, strain tracing, and immunologic characterization together with multi-block analysis of biological, clinical, demographic, and behavioral parameters to investigate LBP effects and correlates of treatment success. LACTIN-V treatment resulted in *L. crispatus*-dominant vaginal bacterial communities in 30% and 35% of recipients at weeks 12 and 24, respectively, representing 3.4- and 4.5-fold higher rates than in placebo recipients. *L. crispatus* colonization among LBP recipients was substantially due to the LACTIN-V strain CTV-05, although dominance by native *L. crispatus* strains increased over time. Microbiota trajectory analysis showed that microbial communities established early frequently persisted. Achieving *Lactobacillus*-dominance ‒ particularly *L. crispatus*-dominance ‒ at early timepoints was strongly linked to persistence of *Lactobacillus*-dominance, while early non-*Lactobacillus*-dominance also tended to be maintained. In exploratory post-hoc analyses, we found that the LBP’s benefits differed based on baseline vaginal microbiota composition, and we identified specific microbial, immune, demographic, and behavioral factors associated with high-level *L. crispatus* colonization among LBP recipients.

To enhance resolution and power, we relied, for most of our analyses, on the observed relative abundances of species, genera, or *de novo* topics rather than on *de novo* clusters or assignment to reference community state types (CSTs). This approach is based on prior work showing that diverse vaginal microbiotas do not exhibit well-defined clusters (Lebeer et al., 2023; Symul et al., 2023) which we also observed in this cohort. We also favored taxa-based reporting to facilitate interpretation and comparison with past or future studies, noting that CST definitions remain in flux as datasets expand beyond predominantly U.S. populations.

To elucidate mechanisms by which the LBP promoted *L. crispatus* colonization, we investigated *L. crispatus* strain dynamics using both metagenomic analysis and cultivation-based strain characterization. Metagenomic strain inferences showed the CTV-05 strain accounted for the majority of *L. crispatus* colonization among LBP recipients, but native *L. crispatus* strains were also observed. Interestingly, among LBP recipients with ascertainable *L. crispatus* strain composition, 96.1% had CTV-05 as their dominant or co-dominant strain at week 4, but this proportion progressively declined, while the number of participants with dominance by one or more native *L. crispatus* strains rose from 3.9% at week 4 to 36.7% at week 24. Recipients with CTV-05 strain dominance at a given visit tended to either maintain CTV-05 dominance or exhibit substantial loss of *L. crispatus* at the ensuing visit. However, recipients with co-dominance of CTV-05 and native strains frequently experienced replacement of CTV-05 by the native strain at the next visit, but very rarely exhibited replacement of the native strain by CTV-05. Further research is needed to determine whether native *L. crispatus* strains that outcompeted CTV-05 have functional characteristics providing selective advantages and whether initial CTV-05 colonization establishes a more permissive environment for eventual transition to native *L. crispatus* strains.

We also examined impacts of MTZ and LBP treatment on mucosal inflammation by measuring vaginal cytokines and chemokines. Significant changes in several cytokines and chemokines were observed in participants for whom treatment successfully shifted microbiota composition to *Lactobacillus-*dominance, and these changes reverted in participants who shifted back to non-*Lactobacillus*-dominance. Most notably, inflammatory cytokines including IL-1α, IL-1ꞵ, and TNFα increased as *Lactobacillus* abundance decreased, whereas IP-10, MIG, and ITAC showed the opposite pattern, consistent with prior reports (Anahtar et al., 2015; Gosmann et al., 2017; Masson et al., 2019, 2014). These effects appeared primarily driven by genus-level *Lactobacillus* abundance rather than a particular species. A prior study examining a subset of LACTIN-V trial participants in whom strain abundance was assessed by qPCR reported that IP-10 levels were lower at week 24 in participants with CTV-05 colonization than those colonized by other *L. crispatus* strains (Armstrong et al., 2022), but we did not observe a similar association in our analysis of the full cohort. Treatment-induced cytokine changes persisted in the LBP arm at week 24 compared to placebo, but this was driven primarily by the LBP’s effect in promoting higher rates of *Lactobacillus*-dominance rather than a strain-specific effect of CTV-05.

Our *post-hoc* analysis of treatment outcomes showed LACTIN-V’s incomplete efficacy in achieving high-level *L. crispatus* colonization and preventing BV recurrence was linked to differences in pre-MTZ microbiota composition. Stratifying participants by their predominant bacterial genus at the pre-MTZ visit revealed between-group differences in achieving *L. crispatus*-dominance at week 24 and in preventing rBV by both weeks 12 and 24. Most prominently, these analyses showed a particular benefit from the LBP in preventing BV recurrence among participants with pre-MTZ *Prevotella*-predominance, whereas participants with pre-MTZ *Ca.* Lachnocurva vaginae (BVAB1)-predominance uniquely exhibited a trend toward higher BV recurrence rates with LBP treatment and little or no benefit from the LBP in achieving *L. crispatus*-dominance. The reasons for these patterns are unclear, but may reflect greater competitive ability or MTZ resistance in *Ca.* Lachnocurva vaginae or its co-occurring species (Alauzet et al., 2010; Aldridge et al., 2001). Since *Ca.* Lachnocurva vaginae remains uncultured (Fredricks et al., 2005; Holm et al., 2020), experimentally testing its antimicrobial susceptibility and ability to compete with other species is currently infeasible, but our results highlight its cultivation and phenotypic characterization as key research priorities. However, we emphasize that these results showing pre-MTZ microbiota effects on LBP efficacy are based on exploratory analyses of a relatively small study cohort, which can introduce arm imbalances through *post-hoc* stratification. They should therefore be regarded as hypothesis-generating observations that require replication and validation in future studies.

Our integrated multiblock analysis identified both expected and unanticipated biological, clinical, demographic, and behavioral correlates of *L. crispatus*-dominant colonization in LBP recipients. Low total bacterial load, low vaginal pH, and low microbiota α-diversity at the post-MTZ visit were associated with *L. crispatus*-dominant colonization, findings that help elucidate microbiota dynamic underlying a *post hoc* analysis of clinical parameters which showed that participants achieving BV cure at the post-MTZ visit had lower rates of recurrence at later timepoints (Hemmerling et al., 2024). Our analysis also showed low pre-MTZ vaginal concentrations of MIG and IP-10 and high concentrations of IL-1β were also associated with *L. crispatus*-dominance at week 4 – an unexpected finding given their opposite correlations with *Lactobacillus* abundance during concurrent visits. The explanation for this observation is unclear, but may involve a more vigorous pre-MTZ mucosal immune response helping to clear BV-associated bacteria, presence of inflammatory but MTZ- or LBP-responsive bacteria in some participants at baseline, or other mechanisms. After week 4, the strongest predictor of a participants’ microbiota category was their microbiota category at the previous timepoint, indicating that microbiota composition established early after LBP treatment best predicts subsequent outcomes. Demographic data including self-declared race and education levels were also linked to microbiota composition at week 4 and beyond, but since race and education were closely correlated and likely associated with unmeasured variables, this precluded clear mechanistic interpretation. Despite evidence for sexual activity as a significant influence on BV and vaginal microbiota composition (Vodstrcil et al., 2025), we did not identify a strong signal for sexual behavior as an explanatory variable for microbiota composition in LBP recipients. This may be because of little variation in self-reported sexual behavior throughout the trial such that effects of protected or unprotected sex may have occurred early and been obscured by one of the many factors associated with early *L. crispatus* colonization. It is also recognized that self-reported sexual activity in clinical trials may not be a reliable measure of actual sexual activity (Jewanraj et al., 2020; Schroder et al., 2003a, 2003b; Zenilman et al., 1995), and reporting biases or inaccuracies can decrease power to detect effects.

In summary, our detailed analysis of the vaginal microbiota effects of a single-strain *L. crispatus* LBP reveals patterns of microbiota composition and strain dynamics underlying the clinical effects of LACTIN-V. We identify key microbiota and host factors associated with treatment success and *L. crispatus* colonization. If validated, these findings could help identify patients most likely to benefit from LBP treatment and guide the discovery of novel therapeutic targets to enhance LBP efficacy and improve women’s health outcomes globally.

### Limitations

This study has several limitations. First, samples from a few participants in the primary clinical trial were not available for microbiome and cytokine analysis (samples were unavailable from 10 of 152 originally reported participants in the LBP arm and 5 of 76 participants in the placebo arm). In addition, technical failures of 16S rRNA gene sequencing precluded microbiota compositional analysis in four additional samples, so our analysis cohort was slightly smaller than the primary trial cohort. Second, the limited size of this Phase 2b study reduced statistical power, especially for heterogeneity and between-arm *L. crispatus* colonization comparisons. Third, the study’s relatively infrequent sampling schedule did not permit high temporal resolution to assess microbiota and strain dynamics. Finally, although detailed data were available regarding sexual, contraceptive, hygiene, and clinical parameters, participants were surveyed on only a narrow range of environmental and sociodemographic variables, precluding detailed investigation of factors that might help explain associations with microbiota composition.

## Methods

### Study design and sample collection

Samples analyzed in this study were obtained as part of a previously reported phase 2b, randomized, double-blind, placebo-controlled trial of the *L. crispatus* LBP LACTIN-V (Osel, Inc., Mountain View, CA) for prevention of rBV (Cohen et al., 2020). Use of samples was approved by the Mass General Brigham Institutional Review Board (IRB Protocol #:2020P002237) as well as the UCSF Institutional Review Board (IRB Protocol#: 19-28337). LACTIN-V is a single-strain LBP formulated as a powder containing a preservation matrix and 2×10^9^ colony-forming units (CFU) per dose of the *L. crispatus* strain CTV-05, which was isolated from a human vaginal sample (Cohen et al., 2020). CTV-05 is administered using a pre-filled vaginal applicator, and was compared in the trial to a placebo formulation consisting of the preservation matrix without CTV-05. The trial was conducted at four centers within the USA and enrolled premenopausal, non-pregnant women aged 18-45 years. Eligibility criteria were previously described (Cohen et al., 2020). Briefly, potential participants attended a screening (pre-MTZ) visit at which they were determined to be eligible for the study if testing revealed presence of BV as determined by both a Nugent score of 4-10 on Gram stain of a vaginal smear (Nugent et al., 1991) and presence of at least three of four Amsel criteria (characteristic vaginal discharge, >20% clue cells on microscopy of a vaginal wet prep, vaginal fluid pH >4.5, and presence of a fishy odor upon addition of 10% potassium hydroxide to a vaginal specimen) (Amsel et al., 1983), as well as negative testing for HIV, syphilis, gonorrhea, chlamydia, trichomonas, and urinary tract infection. Women found to be eligible based on this evaluation completed 5 days of intravaginal MTZ therapy within 30 days of their pre-MTZ visit (**Figure 1A, S1A)**. They then returned to the trial clinic within 48 hours of completing antibiotics (post-MTZ visit) and were randomized in a 2:1 ratio to receive either LBP or placebo after providing written informed consent. The first dose of LBP or placebo was clinician-administered at the randomization (post-MTZ) visit, then doses were vaginally self-delivered daily for the next four days, then twice weekly for ten additional weeks. In-person study visits were scheduled 4, 8, 12, and 24 weeks after randomization, at which vaginal swabs were collected and stored (details below) and clinical report forms (CRFs) were completed. In addition, two phone visits were planned at week 16 and 20 during which a subset of the clinical report forms were filled to capture information on adverse events, menstruation, concomitant medication use, and sexual behavior protected or unprotected by condoms. Participants who desired additional in-person visits were invited to present to the clinics. Swabs and CRFs were collected at these additional visits.

Clinician-collected vaginal swab samples were obtained via speculum exam at in-person study visits. Two types of swabs were collected in parallel for analysis (Cohen et al., 2020). One set of swabs was collected at all in-person visits including the pre-MTZ (screening) visit using the Starplex™ Scientific Multitrans™ Collection and Transportation System (Starplex™ Scientific S1600), which comprises a plastic-shaft Dacron™-tipped swab stored in a glass bead-containing transport medium consisting gelatin (5.0 g/L), sucrose (68.46 g/L), glutamic acid (0.70 g/L), HEPES sodium salt (3.4 g/L), modified Hank’s balanced salts (9.8 g/L), sodium bicarbonate (0.35 g/L), bovine serum albumin (10 g/L) and the antimicrobial agents vancomycin (0.1 g/L), amphotericin B (2.5 mg/L), and colistin (0.015 g/L) at a pH of 7.2-7.8. The other set of swabs was collected at the post-MTZ (randomization) visit and all subsequent visits in the ESwab^Ⓡ^ Liquid Based Collection and Transport System (Copan ESwab 480C^Ⓡ^). Samples were stored at room temperature for 1-4 hours after collection, then frozen at −80°C.

Microbiota sequencing, bacterial isolation, and cytokine analysis was performed on samples stored using the Starplex™ system. Samples were thawed on ice, vortexed at maximum speed for 5 seconds, then the swabs were removed from the transport media and media was divided into aliquots and re-frozen at −80°C. Subsequent processing was performed as described below. Measurement of bacterial load via qPCR was performed using swabs collected via the Copan ESwab^Ⓡ^ system.

### Bacterial isolations and cultivation

Bacterial isolation and cultivation was performed under anaerobic conditions at 37°C in an AS-580 anaerobic chamber (Anaerobe Systems) with an atmosphere of 5% carbon dioxide, 5% hydrogen, and 90% nitrogen (Airgas, Inc.). All culture media was pre-reduced prior to use by being placed in the anaerobic chamber overnight. Bacteria were isolated and cultivated on solid media including *Lactobacillus* MRS agar (Hardy Diagnostics, #G117), Columbia Blood Agar (“CBA”, Hardy Diagnostics, #A16), or CDC Anaerobe Laked Sheep Blood Agar with Kanamycin and Vancomycin (“LKV”, BD BBL™ Prepared Plated Media, #221846). Known *L. crispatus* strains and not-yet-identified isolates obtained on *Lactobacillus* MRS agar were expanded by culture in liquid media consisting of *Lactobacillus* MRS broth (BD #288130) prepared according to manufacturer instructions. Isolates obtained on CBA agar or LKV agar were expanded in liquid media consisting of either Wilkins-Chalgren Anaerobe Broth (Thermo Scientific™ Oxoid™, #CM0643B; prepared according to manufacturer instructions) or of NYCIII broth (American Type Culture Collection (ATCC) medium 1685), whichever produced better growth. NYCIII broth was prepared using a slightly modified version of the standard ATCC protocol (Bloom et al., 2022). Pre-media consisted of 4 g/L HEPES (Fisher Scientific, #BP310-500), 15 g/L proteose peptone no. 3 (BD Biosciences, #BD 211693), and 5 g/L sodium chloride in 875 ml distilled water, which was pH-adjusted to 7.3 and autoclaved on liquid protocol at 121°C for 15 minutes, then cooled and stored at 4°C. One day before use, complete NYCIII broth was prepared from the autoclaved, cooled pre-media by adding dextrose (from a stock of 3 g per 45 ml; Fisher Chemical™, #D16-500) at 7.5% v/v, yeast extract solution (Gibco, #18180-059) at 2.5% v/v, and heat-inactivated horse serum (Gibco, #26050070) at 10% v/v, then sterilized by passage through a 0.22 μm vacuum filter.

To establish a genome sequence for the CTV-05 strain, a cryopreserved pure culture of CTV-05 was obtained from Osel, Inc. The strain was streaked for isolation on *Lactobacillus* MRS agar and cultured for 48 hours, then a single colony was picked into *Lactobacillus* MRS broth and incubated for 20 hours. The broth culture was harvested by centrifugation to obtain bacterial pellets for genomic DNA extraction and sequencing.

Bacterial isolations were performed from twelve selected trial samples (see **Figure 3C-E**, **S3A**) using a modification of previously described methods (Bloom et al., 2022). Since samples had been collected into Starplex™ transport medium containing antibiotics (see above), the samples were thawed on ice, immediately diluted into pre-reduced Dulbecco’s Phosphate Buffered Saline (“PBS”, Millipore Sigma, #D8537) at 1:12 v/v, centrifuged for 10 minutes at 10,000 rcf, then supernatant was removed. Pellets were re-suspended in PBS and re-centrifuged with removal of supernatant two more times, then resuspended in PBS, diluted in serial 10-fold dilutions, and 100 𝜇l aliquots from each dilution were plated evenly on MRS, LKV, and CBA agar in parallel. Plates were incubated for 7 days and multiple examples of each distinct colony morphology from each sample were picked and subcultured onto solid media of the same type as the source media, with an emphasis on colonies from MRS agar with characteristic *L. crispatus* morphology (e.g., **Figure 3D**). After sub-culture for 3-7 days, colonies of the sub-cultured bacteria were picked into MRS broth (for colonies isolated on MRS agar) or into both NYCIII broth and Wilkens-Chalgren broth in parallel and incubated for 1-4 days, depending on growth rate. The resulting liquid cultures were then cryopreserved, with aliquots of each culture centrifuged and pellets saved for genomic DNA extraction and sequencing (see below).

### Nucleic acid extraction for short-read sequencing

Total nucleic acids (TNA) extraction from cervicovaginal swabs for microbiota profiling was performed via a phenol-chloroform method, which includes a previously described bead beating process to disrupt bacteria^70^ and modified for processing in 96-well plate format (Phenol:Chloroform:IAA, 25:24:1, pH 6.6, Invitrogen, #AM9730, which has since been discontinued; Sodium Dodecyl Sulfate 20% Solution, Fisher Scientific, #BP1311-200; EDTA, Invitrogen, #AM9260G; 2-Propanol, Sigma, #I9516-500ML; 3M Sodium Acetate, pH 5.5, Life Technologies, #AM9740). Aliquoted samples were thawed on ice, then TNA extraction was performed using 200uL of well-mixed Star media. The extracted TNA sample was eluted into 80uL of TE buffer (Promega, #V6321).

To extract bacterial genomic DNA (gDNA) for genome sequencing of cultured bacterial strains, each isolated strain was streaked on the indicated solid media and a single, clonal colony was picked into broth culture and incubated in static culture under anaerobic conditions for between 18 and 120 hours (depending on strain growth kinetics). Cultures were centrifuged and gDNA was extracted from the pellets using a plate-based protocol including a bead beating process and combining phenol-chloroform isolation (Anahtar et al., 2016) with QIAamp 96 DNA QIAcube HT kit (Qiagen, # 51331) procedures.

### Bacterial 16S ribosomal RNA (rRNA) gene amplification and sequencing

Bacterial microbiota taxonomic composition in cervicovaginal samples was determined by sequencing the V4 region of the bacterial 16S rRNA gene. The V4 region was amplified via polymerase chain reaction (PCR) using the primer set 515F/806R at 200 pM each (515F primer sequence 5’-AATGATACGGCGACCACCGAGACGTACGTACGGT<u>GTGCCAGCMGCCGCGGTAA</u>-3’ and barcoded 806R primer sequence 5’-CAAGCAGAAGACGGCATACGAGATXXXXXXXXXXXXAGTCAGTCAGCC<u>GGACTACHVGGGTWTCTAAT</u>-3’, in which the underlined sequences in each primer represent the regions of complementarity to 5’ and 3’ ends of the V4 region of the bacterial 16S rRNA gene, respectively, and the barcode positions in the 806R primer are indicated by X; IDT), with the 806R primers barcoded for multiplexing (Bloom et al., 2022; Caporaso et al., 2011). PCR was performed in 25 µl reactions containing 1X Q5 reaction buffer (NEB, #B9027), 0.2 mM of dNTPs (NEB, #N0447), 0.2 μM of each primer, 0.5 unit of Q5 high-fidelity DNA polymerase (NEB, #M0491), and 2 µl of the TNA sample. PCR was performed in triplicate for each sample in the following program: 98°C for 30 seconds, followed by 30 cycles of 98°C for 10 seconds, 60°C for 30 seconds, and 72°C for 20 seconds, followed by a final extension at 72°C for 2 minutes. The triplicate PCR reactions for each sample were combined and amplicon production and size were confirmed on an agarose gel. Negative controls with PCR-quality water (Invitrogen, #10977) as a template were amplified in parallel for each primer barcode mix and assessed in parallel by gel electrophoresis to confirm absence of contamination and non-specific amplification. PCR products were pooled, with the amount for each sample semi-quantitatively adjusted based on its gel band intensity, then purified with a QIAquick PCR purification kit (Qiagen, #28104) and quality controlled with the Qubit^TM^4 Fluorometer (Invitrogen #Q33226), and TapeStation (Agilent Technologies, 4200 TapeStation). Libraries were mixed with 10% PhiX and single-end sequenced on an Illumina MiSeq using a 300-cycle v2 kit (Illumina, #MS-102-2002) employing the custom Earth Microbiome Project sequencing primers (Read 1 sequencing primer sequence: 5’-ACGTACGTACGGTGTGCCAGCMGCCGCGGTAA-3’; read 2 sequencing primer sequence: 5’-ACGTACGTACCCGGACTACHVGGGTWTCTAAT-3’; index sequencing primer sequence: 5’-ATTAGAWACCCBDGTAGTCCGGCTGACTGACT-3’; IDT) (Caporaso et al., 2011). Negative controls for TNA extractions and PCRs were included in each sequencing library. Study samples were sequenced in a total of six libraries. Samples with read counts < 10,000 in initial libraries were re-amplified and re-pooled into subsequent libraries and data from the run producing the highest number of reads for each sample (if the sample was sequenced multiple times) were selected for subsequent analysis. Analyzable 16S rRNA gene data was generated from a total of 1152 out of 1156 available trial samples, with the remaining 4 samples failing due to technical challenges with extraction, amplification, or sequencing.

### Bacterial short-read shotgun whole genome library preparation and sequencing

Shotgun metagenomic and genomic libraries were prepared following a modified protocol of Baym et. al (Baym et al., 2015), using the Nextera DNA Library Preparation Kit (Illumina, #20034211) and KAPA HiFi Library Amplification Kit (Kapa Biosystems, #KK2602). In brief, DNA from each sample was standardized to a concentration of 1ng/mL after quantification with SYBR Green (Invitrogen, #S7653), followed by simultaneous fragmentation and sequencing adaptor incorporation by mixing 1ng of DNA (1 mL) with 1.25 ml TD buffer and 0.25 mL TDE1 provided in the Nextera kit and incubating for 9min at 55°C. Tagmented DNA fragments were amplified in PCR using the KAPA high fidelity library amplification reagents, with Illumina adaptor sequences and sample barcodes incorporated in primers. PCR products were pooled, purified with magnetic beads (MagBio Genomics #AC-60050) and paired-end sequenced on Illumina NovaSeq X with a 300-cycle kit (Psomagen, Inc.).

### Bacterial 16S rRNA gene sequence processing and annotation

Demultiplexing of Illumina MiSeq bacterial 16S rRNA gene sequence data was performed using QIIME 1 version 1.9.188 (Caporaso et al., 2010). Mapping files created in QIIME 1 format were validated using validate_mapping_file.py, then sequences were demultiplexed with split_libraries_fastq.py using parameter store_demultiplexed_fastq and no quality filtering or trimming, and demultiplexed sequences were organized into individual fastq files using split_sequence_file_on_sample_ids.py. Sequence reads were trimmed and filtered using dada2 version 1.6.0 (Callahan et al., 2016), trimming at positions 10 (left) and 230 (right) using the filterAndTrim function with truncQ = 11, MaxEE = 2, and MaxN = 0. Sequences were then inferred, then initial taxonomy assigned using the dada2 assignTaxonomy function with the RDP training database rdp_train_set_16.fa.gz (https://www.mothur.org/wiki/RDP_reference_files). Amplicon sequence variant (ASV) taxonomic assignments were refined via extensive manual review. The resulting annotated sequences were analyzed in R using phyloseq version 1.30.0 (McMurdie and Holmes, 2013) and custom R scripts. Sequence processing and taxonomy assignment was performed blinded to information about participants’ and samples’ clinical and demographic characteristics, treatments, and trial outcomes.

### Total bacterial load via qPCR

Quantification of total bacterial load via qPCR was performed and reported as part of the initial LACTIN-V clinical trial^36^. In brief, DNA extracted from samples stored in the Copan ESwab^Ⓡ^ system were amplified using bacterial 16S rRNA gene primers targeting total bacteria (16S ribosomal DNA, AGAGTTTGATCCTGGCTCAG, GCTGCCTCCCGTAGGAGT, 312bp). Bacterial concentration was calculated using a standard curve based on serial dilutions of the CTV-05 strain as previously described (Cohen et al., 2020).

### Assessing balance between arms at the pre- and post-MTZ visits

Since pre-intervention (*i.e.,* pre-MTZ or post-MTZ) microbiota composition may be associated with differential microbiota composition post-intervention, imbalances between arms could lead to biases in our primary outcome benefit ratio estimates. To assess whether significant imbalances in microbiota composition existed in this cohort, we performed a PERMANOVA analysis, as implemented in the vegan R package, to test whether the intervention arm (LBP *vs* placebo) explained significant variability in microbiota β-diversity, computed using the Bray-Curtis dissimilarity on ASV relative abundances.

### Benefit Ratios

Benefit ratios, and associated confidence intervals and *p*-value were computed using Wald’s method as implemented in the epitools R package (Aragon, 2004).

### Identification of microbiota topics from 16S rRNA gene sequencing count data

Microbiota topics were identified using a modification of a previously described approach (Symul et al., 2023). One main difference is that here, *Lactobacillus* topics (i.e., topics composed exclusively of *Lactobacillus* species) were defined independently from non-*Lactobacillus* topics (*i.e.,* topics composed exclusively of non-*Lactobacillus* species), which were identified by fitting a Latent Dirichlet Allocation model (Blei et al., 2003) to the non-*Lactobacillus* ASV counts aggregated at the species level. This was done to facilitate the interpretation of topic composition and, specifically, to allow for “pure” topics for the most prevalent *Lactobacillus* species in this cohort (*L. crispatus*, *L. iners*, and *L. jensenii*).

To determine *K*, the optimal number of non-*Lactobacillus* topics, we relied on a method called “topic alignment” (Fukuyama et al., 2023) which examines the robustness of topics across resolutions (increasing values of *K*) and provides diagnostics scores that facilitate the identification of spurious topics. We selected *K* to minimize the number of spurious topics (flagged by low coherence scores) and such that the number of paths (collection of similar topics across resolution) in the alignment presented a plateau (Fukuyama et al., 2023) (**Figure S2A**-**B**). The estimated proportions (relative abundances) of non-*Lactobacillus* topics in each sample (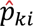 where 𝑘 is the topic and 𝑖 is the sample) were computed by multiplying the proportions estimated by the model on non-*Lactobacillus* counts 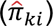 by the total non-*Lactobacillus* proportions in each samples (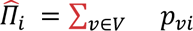 where 𝑝*_vi_* is the observed proportion of ASV 𝑣 in sample 𝑖 and 𝑉 is the set of non-*Lactobacillus* ASVs, such that 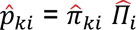).

*Lactobacillus* topics were defined as follows: *Lactobacillus* species which reached 50% of a microbiota composition in at least 10 samples made up their own topic, while the remaining *Lactobacillus* species were grouped into a single topic (“Other *L.*”) as their total prevalence and abundance was overall small (**Figure S2C**). The composition of this topic was estimated from the species average prevalences in this cohort. The estimation of proportions of *Lactobacillus* topics in each sample was straightforward: they were computed from the proportions of the corresponding species in each sample.

### Isolate genome assemblies from short-read sequencing

Paired-end short-read genomic sequencing data from bacterial isolates were processed through a quality control and assembly pipeline. Raw reads were first trimmed for adapter sequences using Cutadapt v4.6 with the Nextera adapter sequence (CTGTCTCTTAT). Quality filtering was then performed using Sickle-trim v1.33 with a quality threshold of Q20 and minimum read length of 50 bp after trimming. Read quality was assessed using FastQC. De novo genome assembly was performed using Unicycler v0.5.0 in standard mode with default parameters. Unicycler produces high-quality assemblies from short reads alone by optimizing SPAdes assembly, followed by graph simplification. The resulting assemblies were annotated using Bakta v1.9.4 with the Bakta database v5, which provides comprehensive bacterial genome annotation including coding sequences, rRNAs, tRNAs, and other genomic features. Assembly completeness and quality were evaluated using BUSCO v5.5 with the Bacteroidales lineage dataset (bacteroidales_odb10), providing a measure of genome completeness based on conserved single-copy orthologs.

### Strain CTV-05 long-read DNA extraction, sequencing, and completed genome assembly

For long-read (Oxford Nanopore Technologies) genome sequencing of the CTV-05 strain, a bacterial pellet was prepared from a clonal colony cultured in *Lactobacillus* MRS broth media as described above. gDNA was extracted using a non-bead-beating protocol to minimize shearing of DNA fragments, with extractions performed and sequenced using five parallel aliquots of the same original broth culture to maximize yield and consistency. Since *L. crispatus* is known to have a thick cell wall containing surface-layer (S-layer) proteins, cell pellets were resuspended with 5M lithium chloride (Molecular Dimensions #MD2-100-43) and then washed with PBS to begin to degrade the cell wall. Using the manufacturer’s protocol for the MasterPureTM Gram Positive DNA Purification Kit (Biosearch Technologies #MGP04100), the bacterial cells were then lysed, proteins and RNA were digested, and genomic DNA (gDNA) was extracted. Extracted gDNA concentration was diluted 1:5 and measured using the Qubit^TM^ 4 Fluorometer, then DNA libraries were prepared using Oxford Nanopore Sequencing’s Rapid Barcoding Kit (Oxford Nanopore Technologies #SQK-RBK004; note that this product has been discontinued as of March 2024). In brief, 400 ng of gDNA from five replicate extractions was barcoded, pooled, purified, and then loaded onto the MinION Flow Cell using R9.4.1 chemistry (Oxford Nanopore Technologies #FLO-MIN106D; note that this product has been discontinued as of July 2024) in the MinION Sequencing Device (Oxford Nanopore Technologies #MIN-101B). To produce the maximum number of reads possible using the Rapid Barcoding Kit, the sequencing run was continued for a full 72 hours.

Demultiplexed genomic long-reads from MinKNOW v2.2 were concatenated, and reads were separately filtered for quality and length using Filtlong v0.2.1 (https://github.com/rrwick/Filtlong). Filtered reads were subsampled to make 12 different read sets using Trycycler v0.5.3 (Wick et al., 2021) and duplicate reads from each read set were removed using BBMap v39.00 (Bushnell, 2014). The read sets were assembled using Flye v2.9.1 (Kolmogorov et al., 2019), Minipolish v0.1.3 (Wick and Holt, 2021), Raven v1.8.1 (Vaser and Šikić, 2021), and Canu v2.2 (Koren et al., 2017). Contigs from different assembly methods were clustered based on similarity to generate consensus sequences and improve assembly quality using Trycycler. Clusters were manually chosen and Trycycler was used to reconcile the contigs within each cluster. Multiple sequence alignment and read partitioning were performed on the reconciled contigs using Trycyler to understand sequence variation within each cluster and assign each read to the cluster it best aligned with respectively. Trycycler was then used to generate a consensus contig sequence for each cluster based on the multiple sequence alignment and read partitioning steps. Medaka v1.7.2 (https://github.com/nanoporetech/medaka), a tool for creating consensus sequences specifically from nanopore sequence data, was used to polish the Trycycler consensus sequences. The consensus sequences were then combined, after which Polypolish v0.5.0 (Wick and Holt, 2022) was used to polish the long-read sequence with Illumina short-read data generated from an aliquot of the same clonal culture of CTV-05 (extracted and sequenced as described above). The complete CTV-05 genome was annotated using Bakta v1.7.0 (Schwengers et al., 2021) and published online as part of this manuscript under NCBI BioProject reference number: PRJNA1303956.

### Construction of *L. crispatus* core SNP database for strain analysis

A GT-Pro database of biallelic core sites was constructed using 236 *L. crispatus* genomes using k-mers with thresholds for SNP prevalence of 0.95 and minor allele frequencies of 0.02 with MAAST (Shi et al., 2023, 2022; Shih et al., 2025). Then k-mers for samples were counted to generate a metagenotype, or a matrix of the number of reads at biallelic SNP sites for a single species (Smith et al., 2022). This matrix was then used by StrainFacts to infer suitable allele and relative abundance matrices that represent strains across all given samples (Shih et al., 2025).

### *L. crispatus* strain estimation in metagenomes

Raw metagenomic paired-end reads from vaginal swab samples were quality-trimmed and adapter-filtered using Cutadapt (v3.9) (Martin, 2011). Human reads were depleted by aligning reads to the T2T-CHM13 reference genome using HISAT2 (v2.2.1) (Kim et al., 2019). Processed reads were profiled using GT-Pro (v1.0.1) against the custom SNP database (Shi et al., 2022). Resulting metagenotype profiles were analyzed with StrainFacts (v0.6.0) to infer strain genotypes and abundances for *L. crispatus*, excluding samples with <25% coverage of the biallelic SNPs (Smith et al., 2022). The fit was then run through a cleanup step to remove low abundance strains and strains with extremely similar genotypes using sfacts cleanup_fit −-abundance 0.01 −-dissimilarity 0.01 −-discretized. Analysis was performed on samples with ≥5% overall relative abundance of *L. crispatus* as determined by 16S rRNA gene sequencing based on evaluations of parameters supporting accurate StrainFacts LBP strain inference (Shih et al., 2025). *L. crispatus* strain inference was successful in 313 of 343 samples in which species relative abundance exceeded the analysis threshold, while strain analysis was unsuccessful in the remaining 30 samples due to failed or inadequately deep shotgun metagenomic sequencing. A 10% strain proportional abundance was used as a threshold for confident strain identification based on prior simulations and experimental validation (Shih et al., 2025).

### Strain analysis of isolate genomes and comparison to metagenomically inferred strain genotypes

Isolate genomes were taxonomically identified using GTDB-Tk v2 (Chaumeil et al., 2022). Gene-calling was performed for *L. crispatus* isolate genomes, including the completed CTV-05 genome (see above) using Prodigal (Hyatt et al., 2010). Core ribosomal proteins were identified using HMMER (Eddy, 2023), aligned using mafft (Katoh et al., 2019; Kuraku et al., 2013), then concatenated and a phylogenetic tree was constructed using fasttree (Price et al., 2010, 2009), which was used to compute branch lengths to identify isolates representing distinct strains and determine which isolates represented CTV-05 or endogenous strains. GT-Pro (Shi et al., 2022) was then used to calculate genotypes from representative isolate reads for comparison to metagenomically inferred strain genotypes, calculated based on Jaccard similarity of the StrainFacts genotypes (Shih et al., 2025).

### Measurement of vaginal mucosal cytokines and chemokines

Cytokines and chemokines were measured using a previously described custom 20-plex High Sensitivity Luminex Assay Kit from EMD Millipore that measures interferon gamma-induced protein (IP-10), interleukin 8 (IL-8), interleukin 6 (IL-6), monokine induced by interferon gamma (MIG), interferon-inducible T cell alpha chemoattractant (ITAC), interleukin 1 alpha (IL-1α), interleukin 1 beta (IL-1β), macrophage inflammatory protein-1 alpha (MIP-1α), macrophage inflammatory protein-1 beta (MIP-1β), macrophage inflammatory protein-3 alpha (MIP-3α), tumour necrosis factor alpha (TNFα), interleukin 21 (IL-21), interleukin 17 (IL-17), interferon gamma (IFNγ), interleukin 23 (IL-23), interleukin 12 (IL-12 p70), interleukin 13 (IL-13), interleukin 10 (IL-10), interleukin 4 (IL-4), and interleukin 5 (IL-5) (Gosmann et al., 2017). Reagents were used as supplied by the manufacturer, except the Mixed Beads solution, detection antibody mixture, and Streptavidin-phycoerythrin (Strep-PE) were diluted 3-fold (1:2 volume:volume ratio) for use. The Mixed Beads were diluted in Bead Diluent, while the detection antibodies and Strep-PE were diluted individually in Assay Buffer. All reagents were from a single manufacturer lot to minimize batch effects.

Samples were first pre-processed to remove interfering mucus and debris. In brief, 200 𝜇L aliquots of each vaginal swab supernatant were thawed on ice, vortexed for 10 seconds to resuspend, then centrifuged at 1000 rcf for 15 minutes at 4°C to pellet mucous and cells. The maximum amount of supernatant recoverable from each sample without disturbing the resulting pellet was then transferred to a well of a 0.22 m PVDF filter plate (EMD Millipore #MSGVS2210), filtered by centrifugation at 2,451 rcf for 1 hour at 4°C, then transferred to freezer-safe tubes and stored at −80°C until assayed. Samples were assayed in 96-well plates as per manufacturer protocol with the addition of periodic sonication steps to minimize bead clumping. Each plate assay included one blank background control, 7 standard serial dilutions assayed in duplicate, two manufacturer-supplied Quality Control (QC) samples assayed in duplicate, 72 experimental samples, and 4 biological control samples used as internal quality controls on all plates. Batches of standards and QC samples were prepared from the kit on the day of the assay according to manufacturer protocol. For each sample or standard, 25 𝜇L each of Assay Buffer, Mixed Beads, and sample were aliquoted into each plate well. After the addition of Mixed Beads, all incubation steps were performed while the plate was protected from light. Plates were sonicated for 30 seconds at room temperature in a bath sonicator, then incubated for 16-18 hours on a horizontal plate shaker at 600 rpm in 4°C. An additional 30 second sonication at room temperature was then performed. A handheld plate magnet was applied to the base of the plate to retain the magnetic beads and excess sample and reagents were removed via three consecutive washes using manufacturer-supplied 1X Wash Buffer. Detection antibodies were added as per manufacturer protocol and plates were incubated for 1 hour at room temperature on a horizontal plate shaker at 600 rpm. Next, Strep-PE reagent was added, followed by a 30 minute incubation at room temperature on a horizontal plate shaker at 600 rpm. Plates were washed 3 times as above using the magnet. After the final wash, 150 𝜇L of sheath fluid was added to all wells and a final 30 second sonication at room temperature was performed. Analyte concentrations were measured using a FLEXMAP 3D instrument with Luminex xPONENT software.

### Cytokine and chemokine concentration transformations

Cytokine and chemokine concentrations were log-transformed (log) to stabilize the mean-variance relationship. Compounds with concentration values below the lower limit of quantification (LLOQ) were imputed at half the LLOQ; those with concentration values above the upper limit of quantification (ULOQ) were imputed at the ULOQ (**Figure S4A**).

Measurement of soluble molecules in swab samples can be complicated by a “size effect” phenomenon whereby a component of between-swab variation in concentrations is due to technical differences in the amount of material collected on each swab. In such situations, the first principal component (PC1), which reflects the “size” of observations (Jolicoeur and Mosimann, 1960), can primarily be driven by the amount of material on the swab rather than biological variation. Consistent with this size effect phenomenon, we observed that per-sample cytokine/chemokine concentrations were highly collinear, with PC1 of the standardized log-transformed cytokine concentrations accounting for over 50% of the total variance (**Figure S4B**) but showed no substantial correlation with clinical and biological factors such as proportions of total *Lactobacillus* or of *Lactobacillus crispatus*, or participants’ contraceptives (**SI**). Such size effects can be addressed by subtracting PC1 from the data. We therefore performed PC1 subtraction by re-assigning scores corresponding to the 1st PC to a value of 0, then transforming the data back to its original variable space using the transposed rotation matrix (**SI**). Sensitivity analyses were performed to assess the impact of this transformation on findings (**SI**). Finally, due to the large uncertainty regarding their distributions, cytokines with values below the LLOQ in ≥60% of the samples or with values above the ULOQ in ≥30% of the samples were excluded from the analysis (**Figure S4A**).

### Associations between microbiota composition and cytokine profiles

To quantify the association between microbiota composition and cytokine profiles, we computed the RV coefficient between the microbiota composition as expressed as topic proportions and cytokine transformed log_10_-concentrations. Associated *p*-value was computed with a permutation test (Bougeard and Dray, 2018; Dray and Dufour, 2007). To confirm that correlation between these tables was not driven by potential subject (longitudinal) effects, we also computed correlation between the two tables at each visit independently. If correlations were found significant, further analyses were carried out to characterize the relationship between tables. Specifically, DISTATIS (Abdi et al., 2012, 2005), as implemented in the distatisR R package, was used. This method presents the advantage of estimating compromise scores (and partial residual scores) directly from dissimilarity matrices such that ecological measures of (dis)similarity, such as the Bray-Curtis dissimilarity, can be used for representing microbiota composition. Associations with the original variables can then be assessed using correlations and displayed in correlation circles as typically done with PCA, PCoA, or NMDS results. The minimum number of components used for display was chosen based on the presence of an elbow in the DiSTATIS scree plot.

### Estimating effect heterogeneity

Binary outcomes (Y: ≥50% colonization by *L. crispatus* at week 12 or 24 or absence of rBV by week 12 or 24) were predicted using logistic regression (logit link function) with input variables representing the intervention arm (*A*: LBP or placebo), the baseline (pre-MTZ) microbiota (*V*: stratified by most prevalent genus, CST, or topic proportions), and their interactions (*A:V*). In stratification analyses, strata with less than two participants per arm were excluded from the analysis and visualizations. Since *Lactobacillus* species other than *L. iners* had low pre-MTZ relative abundance, prior to determining fits relying on topic proportions, *Lactobacillus*-dominated topics were agglomerated such that the proportion of total *Lactobacillus* was considered, which improved fit convergence. The statistical significance of effect heterogeneity was tested by comparing the null model (which only includes the intervention arm as predictor) with the full model where pre-MTZ microbiota and interaction effects were included (analysis of deviance F test). *P*-values were adjusted to account for multiple hypotheses testing using the Benjamini-Hochberg procedure (Benjamini and Hochberg, 1995). For the stratified analyses, CI were computed using Wilson scores due to the small sample size in each stratum. When pre-MTZ microbiota composition was included as a quantitative multivariate variable, counterfactual probabilities of success and associated 95% confidence intervals were predicted from the logistic regression fitted model by setting the intervention arm to LBP or placebo. The predicted participant-level odd ratios were computed as the ratio between predicted odds of *L. crispatus* colonization at week 12 and 24 or rBV by week 12 or 24 had participants been receiving LBP or the placebo.

### Identifying factors associated with successful *L. crispatus* colonization in intervention arm

To identify demographic, clinical, behavioral, or microbiologic factors associated with successful *L. crispatus* colonization (≥ 50%) in the intervention arm, we relied on a multiblock partial least square discriminant analysis (MB-PLS-DA) (Brandolini-Bunlon et al., 2019) where the 3-category response variable indicated whether (1) relative abundance of *L. crispatus* was ≥50%, (2) relative abundance of total *Lactobacillus* (any species) was ≥50% but relative abundance of *L. crispatus* was <50%, or (3) the relative abundance of total *Lactobacillus* was <50%. MB-PLS-DA relies on the same principles as PLS-DA but allows for explanatory variables to be grouped into several thematic blocks, which enables the computation of block importance indices and covariances with the response block (Bougeard et al., 2011; Brandolini-Bunlon et al., 2019). The description of each block and associated variables is provided in Table S6.

Several explanatory variables were correlated. For example, the abundances of some cytokines correlated with microbiota composition (**Figure 4**), and the baseline α-diversity was lower in White participants (**Figure S7C**, p-value < 0.05). We addressed these existing correlations between explanatory blocks and/or variables in two different ways depending on our assumptions on the underlying correlation source or cause. We either relied on nested models to evaluate the additive predictive power of specific blocks (*e.g.,* demographics) that had variables correlated with microbiological blocks or used one variable or one block to predict the values of another one and included the residuals instead of the observed values in the model. This was indicated by the symbol (r) in the variable or block names (see **SI** for details). We used this approach for the cytokine blocks whose residuals were computed using their PLS-predicted abundances based on microbiota composition (**SI**). Similarly, we computed residual microbiota composition, α-diversity, and pH at the previous visit such that topic proportions, α-diversity, or pH were relative to those expected based on the participants’ colonization status at the same visit.

Categorical variables such as race or birth control were included using one-hot encoding, and variables were standardized. To ensure convergence of the fits in cross-validation or using the bootstrap, we added Gaussian noise with very small variance (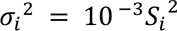 where 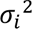 is the variance of the Gaussian noise for variable 𝑖, and 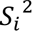 the empirical variance of the 𝑖^th^ variable) to one-hot encoded variables that had few participants in some categories.

Given that our categorical response had three categories, we selected two latent components for the MB-PLS-DA models to avoid overfitting. Further, we performed cross-validation analyses (40 random 75-25% split into calibration and validation sets) and found that two latent components maximized the mean average F1 score on the validation sets for all models except for the initial phase of the placebo in which no variables were found to predict our response, leading to poor and unstable performances in cross-validation (**Figure S6B** and **S8A**). This was consistent with the presence of an elbow in the screeplots and robust to other choices of metrics such as accuracy or RMSE (**SI**). Relative cumulative block importance indices were computed by dividing cumulative block importance, the BIPC as in (Brandolini-Bunlon et al., 2019), by the total inertia of each block, which corresponds to the expected BIPC if all variables had a similar importance (**SI**).

## Data Availability

Sequencing data and genome assemblies generated as part of this study are posted under NCBI BioProject reference number: PRJNA1303956. Analysis code are available on our publicly available GitHub repository (https://github.com/lasy/IIb-LACTIN-V-omics-reanalysis/).

## Acknowledgments

We thank members of the Vaginal Microbiome Research Consortium for helpful discussions. Funding was obtained by D.S.K from the Gates Foundation (INV-033690). S.M.B. was partially supported by NIH grant 1K08AI171166.

## Author Contributions

D.S.K., S.P.H., S.M.B., and L.S. conceptualized the research with input from J.E., C.M.M., A.H., and C.R.H.; D.S.K. and S.P.H. supervised the project; S.M.B., L.S., J.E., J.X., S.H., J.S., and S.P.H. developed and validated methods and models; S.M.B., J.X., S.H., A.S., C.M.M., T.P.P., A.K., and F.A.H. generated samples and performed experiments; L.S., S.M.B., J.E., J.S., A.H., A.K., F.A.H., and S.P.H. performed data analysis; L.S., S.M.B., and D.S.K. led writing of the manuscript with contributions from S.P.H., J.E., J.X., S.H., C.M.M., A.H., T.P.P., A.K., F.A.H., and C.R.H.; All authors provided critical feedback on methods, results, analysis, and writing.

## Data and code availability

Sequencing data and genome assemblies generated as part of this study are posted under NCBI BioProject reference number: PRJNA1303956. Analysis scripts and SI are available on our publicly available GitHub repository (https://github.com/lasy/IIb-LACTIN-V-omics-reanalysis/).

## Supplementary Tables and Figures

**Table S1.**
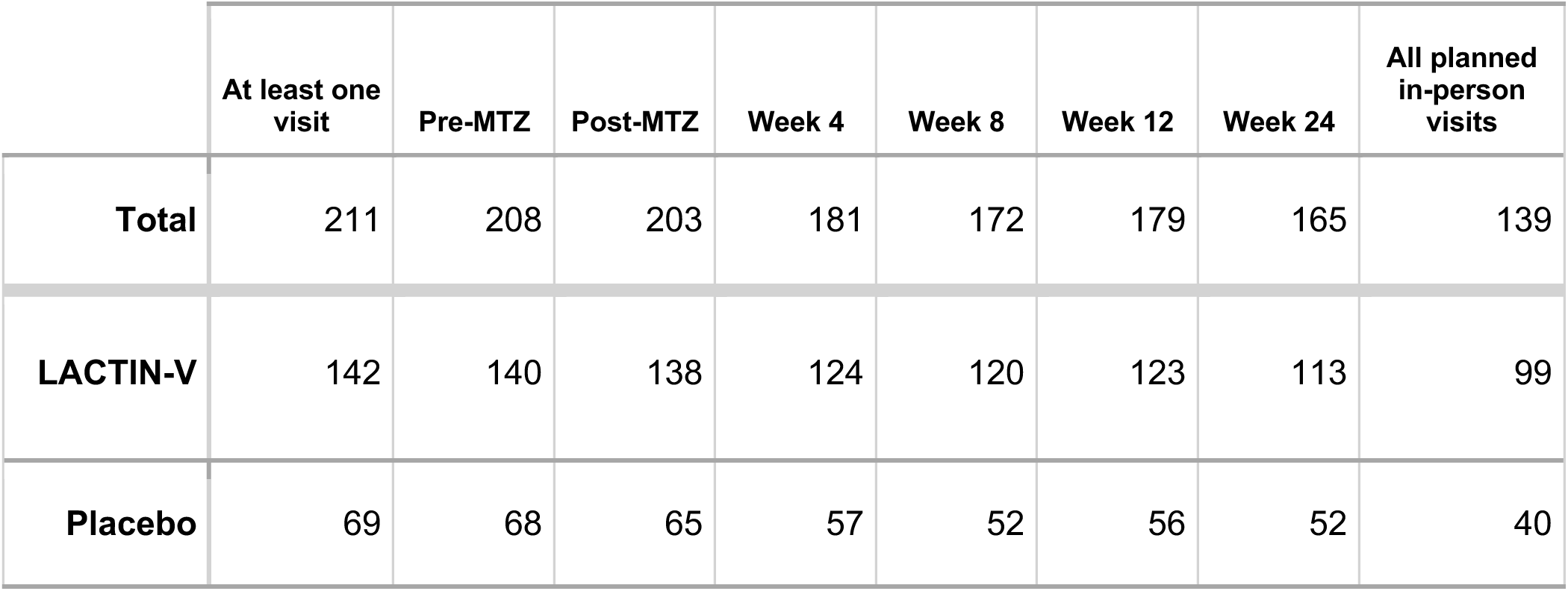
Number of participants in each study arm (rows) with available 16S rRNA gene sequencing data at any, each, or all planned in-person visits (unscheduled clinic visits not shown).

|  | At least one visit | Pre-MTZ | Post-MTZ | Week 4 | Week 8 | Week 12 | Week 24 | All planned in-person visits |
| --- | --- | --- | --- | --- | --- | --- | --- | --- |
| Total | 211 | 208 | 203 | 181 | 172 | 179 | 165 | 139 |
| LACTIN-V | 142 | 140 | 138 | 124 | 120 | 123 | 113 | 99 |
| Placebo | 69 | 68 | 65 | 57 | 52 | 56 | 52 | 40 |

**Table S2.** At the post-MTZ visit, number and proportion of samples dominated by one of the top *Lactobacillus* species (top 3 rows), a mixture of *Lactobacillus* (4th row), or by non-*Lactobacillus* species in both arms (2nd column), the LACTIN-V arm (3rd column), or the placebo arm (4th column).

|  | % (n) [both arm] | % (n) [LACTIN-V] | % (n) [Placebo] |
| --- | --- | --- | --- |
| ≥ 50% *L. iners* | 52 % (106) | 52 % (72) | 52 % (34) |
| ≥ 50% *L. jensenii* | 8 % (16) | 9 % (13) | 5 % (3) |
| ≥ 50% *L. crispatus* | 1 % (2) | 1 % (1) | 2 % (1) |
| ≥ 50% *Lactobacillus* (no individual species ≥ 50%) | 13 % (27) | 14 % (20) | 11 % (7) |
| < 50% *Lactobacillus* | 26 % (52) | 23 % (32) | 31 % (20) |

**Table S3.**
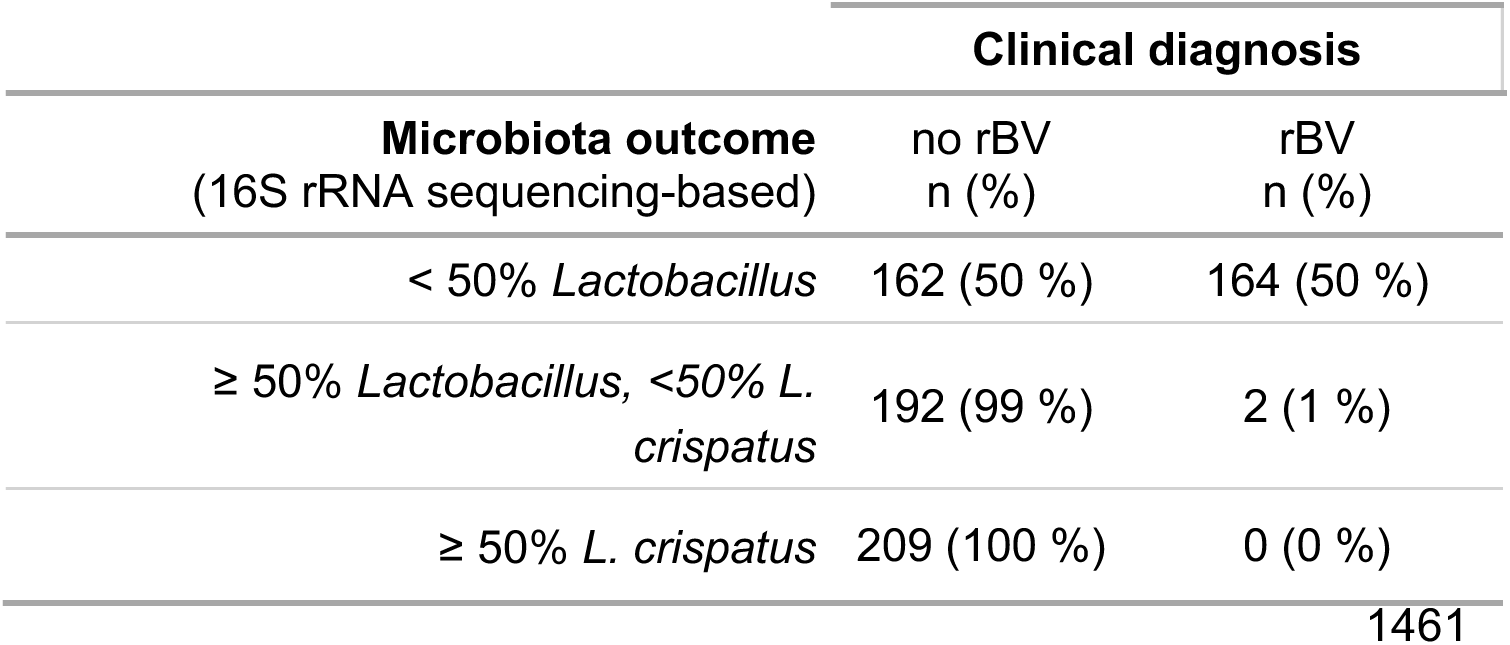
Number of visits (n) at which participants did not (left column) or did (right column) meet diagnostic criteria for recurrent BV (rBV). Participants are shown according to microbiota category. Percentages sum to 100% for each category.

**Table S4.** Contingency table showing the correspondence between Week 4 and Week 12 microbiota categories for LACTIN-V recipients with available data from both visits.

| Category at week 4 | Category at week 12 |  |  | Total |
| --- | --- | --- | --- | --- |
|  | ≥ 50% *L. crispatus* | ≥ 50% *Lactobacillus*,<br>< 50% *L. crispatus* | < 50% *Lactobacillus* |  |
| ≥ 50% *L. crispatus* | 25 | 14 | 12 | 51 |
| ≥ 50% *Lactobacillus*,<br>< 50% *L. crispatus* | 6 | 12 | 8 | 26 |
| < 50% *Lactobacillus* | 4 | 4 | 30 | 38 |
| Total | 35 | 30 | 50 | 115 |

**Table S5.** Contingency table showing the correspondence between Week 4 and Week 24 microbiota categories for LACTIN-V recipients with available data from both visits.

| Category at week 4 | Category at week 24 |  |  | Total |
| --- | --- | --- | --- | --- |
|  | ≥ 50% *L. crispatus* | ≥ 50% *Lactobacillus*,<br>< 50% *L. crispatus* | < 50% *Lactobacillus* |  |
| ≥ 50% *L. crispatus* | 27 | 9 | 11 | 47 |
| ≥ 50% *Lactobacillus*,<br>< 50% *L. crispatus* | 6 | 6 | 11 | 23 |
| < 50% *Lactobacillus* | 5 | 10 | 22 | 37 |
| Total | 38 | 25 | 44 | 107 |

**Table S6.**

| Block | Variable | Description |
| --- | --- | --- |
| Demographics | Age | Age of the participant at enrollment. |
|  | N past BV | Number of past BV episodes (coded as a number on the following scale: (1) None, (2) 1-2, (3) Unknown, (4) 3-4, (5) 5 or more). |
|  | Education level | Education level of the participant (coded as a number on the following scale: (1) Did not complete high school, (2) Completed high school, (3) Completed junior college, (4) Completed college (undergraduate degree), (5) Completed graduate degree). |
|  | Race | Self-declared race of the participant (Categories with few participants were merged into "Other"). |
| Vag. env. pre-MTZ | $\alpha$ diversity | Shannon diversity index of baseline (pre-MTZ) microbiota, computed on taxa proportions quantified from 16S rRNA gene sequencing data. |
|  | pH | Vaginal pH at the pre-MTZ visit, measured at study sites. |
| Microbiota pre-MTZ |  | Pre-MTZ microbiota composition described as topic proportions. |
| Cytokine (r) pre-MTZ |  | Pre-MTZ cytokine residual levels. These values are computed as the difference between the transformed cytokine/chemokines levels and their predicted value based on microbiota composition expressed as topic proportions. |
| Coloniz. cat. prev. v. | | Colonization category at the previous visit. One of these mutually exclusive categories: $\geq 50\%$ *L. crispatus*, $\geq 50\%$ *Lactobacillus* and $< 50\%$ *L. crispatus*, or $< 50\%$ *Lactobacillus*. |
| Vag. env. prev. v. | $\alpha$ diversity (r) | Difference between the Shannon diversity index at the previous visit (w.r.t the visit of the response variable), computed on taxa proportions quantified from 16S rRNA gene sequencing data, and its expected value based on colonization category at the same visit. |
|  | pH (r) | Difference between vaginal pH at the previous visit, measured at study sites, and its expected value based on colonization category at the same visit. |
|  | log10(bact. load) | Log<sub>10</sub> total bacterial load at the previous visit, quantified by qPCR. |
| Microbiota (r) prev. v. |  | Difference between the observed microbiota composition at the previous visit described as topic proportions and the average microbiota composition within each colonization category. |
| Cytokines (r) prev. v. |  | Cytokine residual levels at the previous visit. These values are computed as the difference between the transformed cytokine/chemokines levels and their predicted value based on microbiota composition expressed as topic proportions. |
| Birth control | Non-hormonal | A binary variable indicating whether participants reported exclusively using one of the following non-hormonal birth control methods including abstinence, use of condoms, or fertility awareness methods. |
|  | IUD (Hormonal) | Same as above for hormonal intra-uterine device. |
|  | IUD (Non-hormonal) | Same as above for non-hormonal intra-uterine device (copper IUD). |
|  | Combined | A binary variable indicating whether participants reported using a combination of hormonal contraceptives or switched contraceptive method between the last visit and this one. |
|  | P only | A binary variable indicating whether participants reported exclusively using progestin-only hormonal contraceptive, including the pill, patches, and implants. |
|  | unknown | A binary variable indicating whether a participant's birth control is unknown (missing data). Each participant belongs to only one of each birth control category. |
| Sexual behavior | N new partners | Number of new sexual partners since the last visit. |
|  | Any condomless sex | A binary flag indicating whether the participant reported any sexual intercourse without condoms since the last visit. |
|  | Any condom sex | A binary flag indicating whether the participant reported any sexual intercourse with condoms since the last visit. |
| Perturbations | N douching | Number of days participant reported any douching episode since the last visit. |
|  | N bleeding | Number of days participant reported vaginal bleeding since the last visit. |
| Antibiotics | N vag. abx | Number of days participant reported using vaginal antibiotics since the previous visit. |
|  | N oral abx | Number of days participant reported taking oral antibiotics since the previous visit. |
| Adherence | N missed doses | Difference between the number of LACTIN-V doses that should have been taken between the previous visit and the current visit if the study protocol had been strictly followed and the actual number of doses that were taken. |
|  | Days since last dose | Number of days since the most recent LACTIN-V dose was taken. |

**Figure S1.**
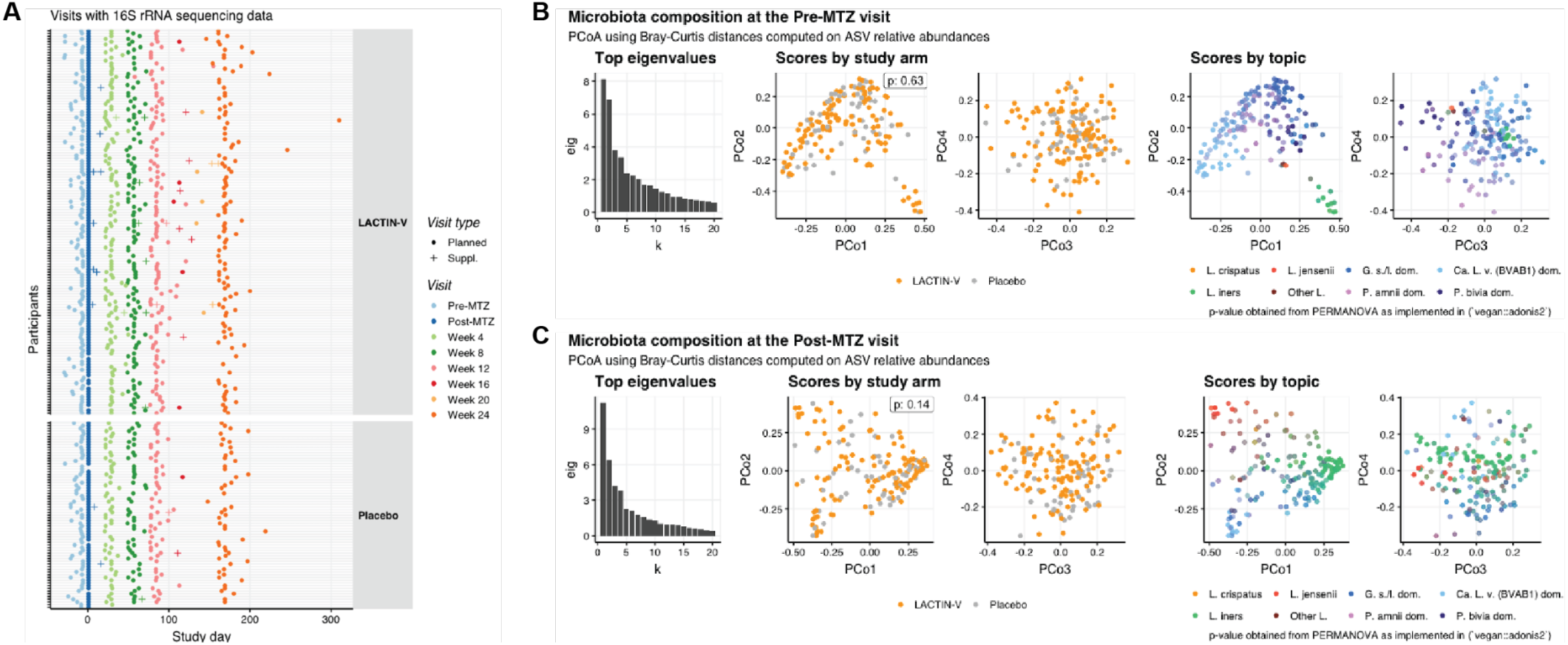
A. Time-line of collected swabs from which 16S rRNA gene sequencing data could be generated. B. Balance between treatment arms in terms of microbiota composition at the pre-MTZ visit. The left panel shows the top 20 eigenvalues of the PCoA (Principal Coordinate Analysis) performed on the Bray-Curtis dissimilarity matrix, computed from the relative abundances of ASVs. The next two panels show the PCoA scores (projection of the samples, each dot is a sample) colored by the intervention arm for the first 4 principal coordinates. The *p*-value in the upper right corner corresponds to the PERMANOVA test on the study arm. The right panels show the same projections as the middle panels, but samples are here colored by topic relative abundance (Figure 2A-B). C. Balance between treatment arms in terms of microbiota composition at the post-MTZ visit, labeled and arranged as for **Figure S1B**.

**Figure S2.**
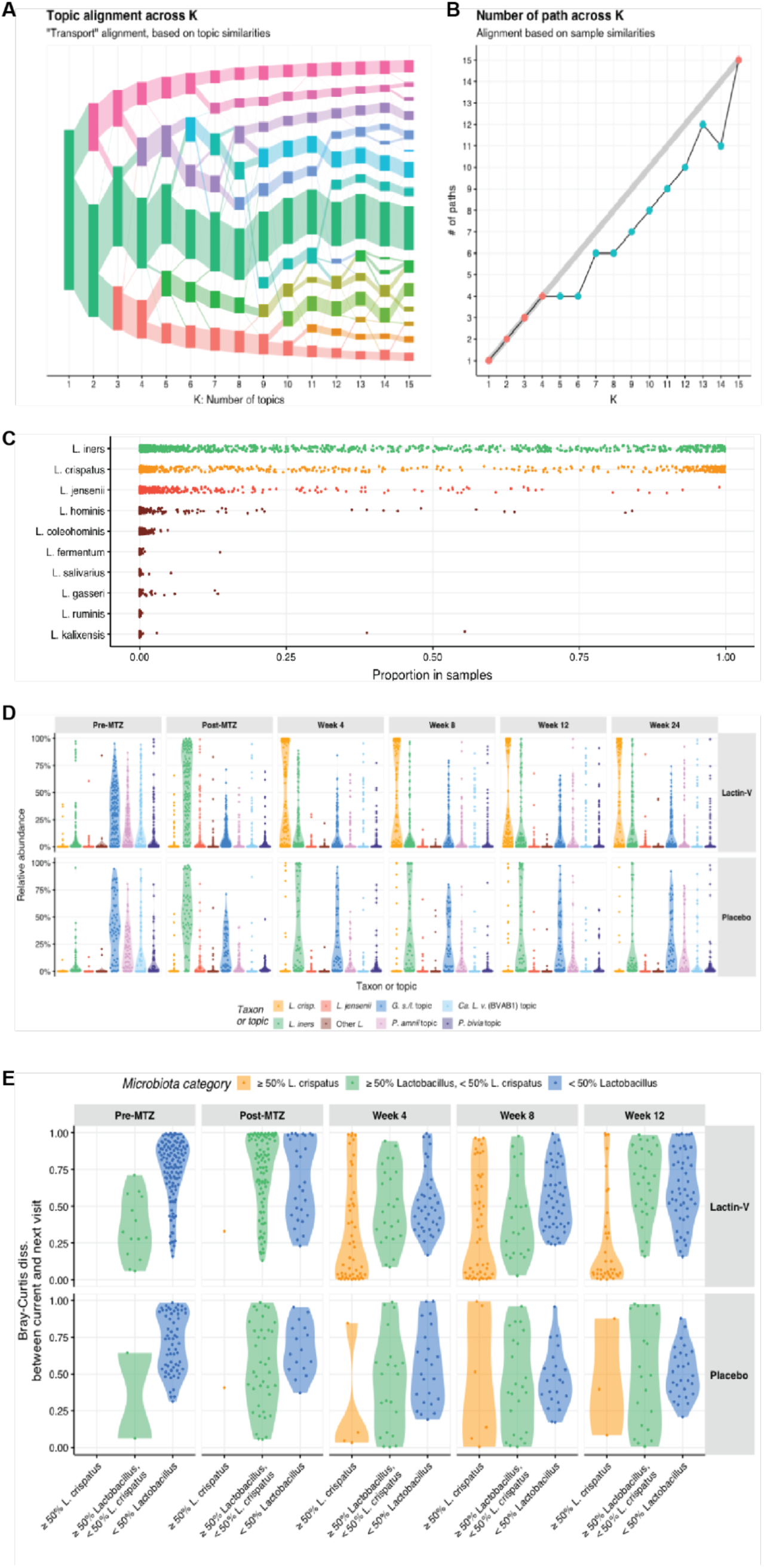
A. “Transport” alignment between topics identified by LDA models fitted on the non-*Lactobacillus* taxa counts for varying total number of topics (K, x-axis). Each rectangle represents a topic; the height of that rectangle represents the summed prevalence of that topic in the population, and the size of the connection between topics is proportional to the degree of similarity between the composition of two given topics. B. The number of paths (y-axis) identified at each resolution (K, the number of topics considered by the LDA models, x-axis) from the “product” alignment of topics; for details see (Fukuyama et al., 2023). A path identifies a series of topics with high alignment scores throughout resolutions. A plateau in the number of paths (here at K = 4) suggests that the 4-topic model is the model that identifies most robust topics (i.e., topics that remain similar across resolution despite the introduction of new, potentially spurious, topics). C. Proportion (x-axis) of each *Lactobacillus* species (y-axis) detected across all samples. Species with a relative abundance > 50% in at least 10 samples made up their own topic. D. Relative abundance of taxon or topic (x-axis, colors) in each arm at each visit. E. Microbiota stability by arm, visit, and microbiota category. Stability is quantified using the Bray-Curtis dissimilarity, described as relative abundances at the ASV level at the participant’s current visit (vertical panels) and subsequent scheduled visit. At each visit, participants are grouped into one of the three categories (colors, x-axis) defined earlier (*L. crispatus*-dominant, *Lactobacillus*-dominant but low-level *L. crispatus*, and low-level *Lactobacillus*).

**Figure S3.**
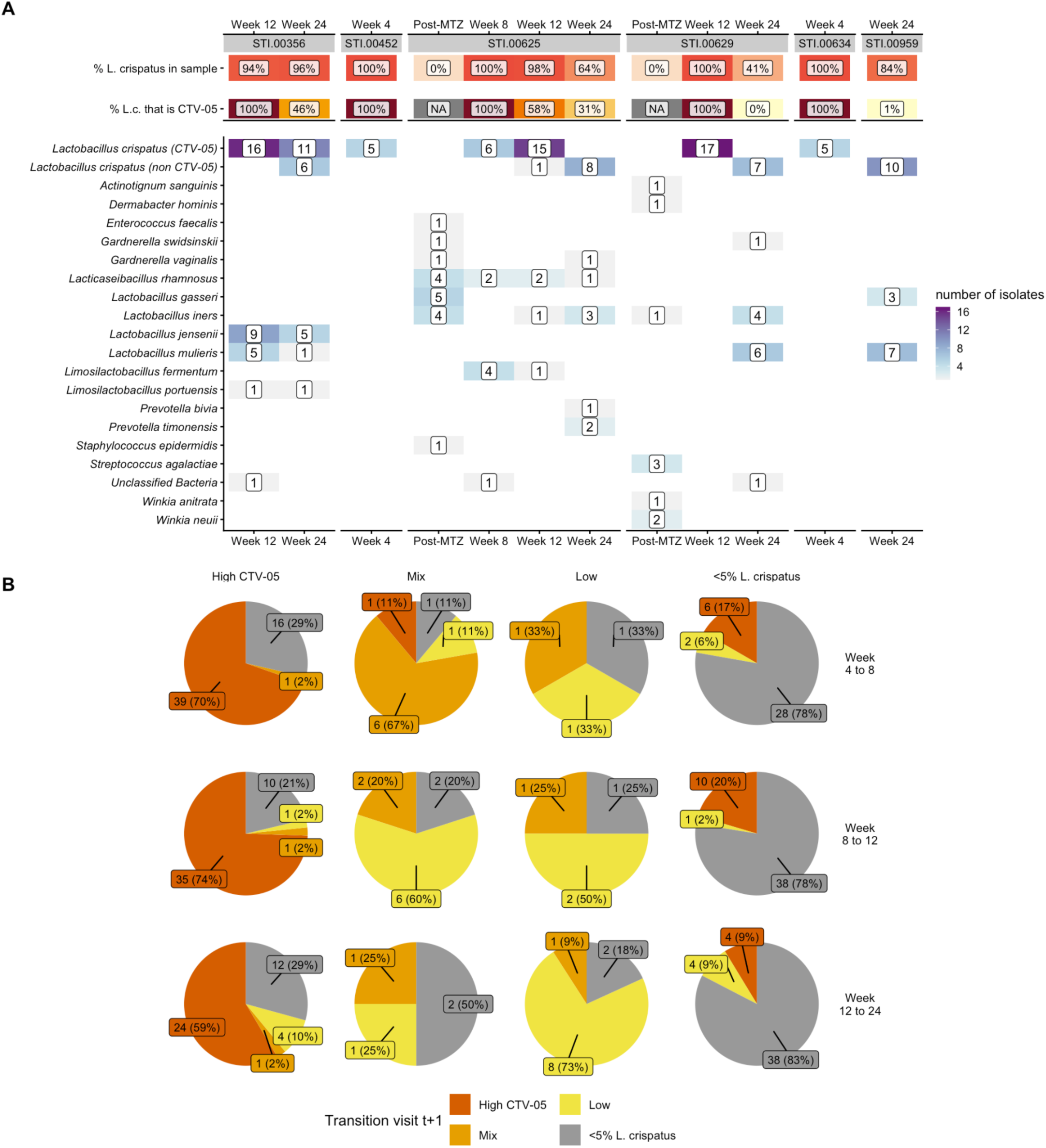
A. Results of targeted isolations from LACTIN-V trial samples, showing source participant, visit number, sample *L. crispatus* relative abundance as estimated by 16S rRNA gene sequencing, CTV-05 proportional strain abundance, and the numbers and taxonomic assignments of genome-sequenced isolates from each sample. *L. crispatus* isolates are separated by whether genome sequencing showed them to be CTV-05 or non-CTV-05 (native) strains. Multiple isolates of genotypically identical strains (including of *L. crispatus* isolates) were obtained from some samples. B. Pie charts for each Visit t strain category from Figure 3F summarizing *L. crispatus* strain category frequencies at Visit t+1, shown individually for each Visit t.

**Figure S4.**
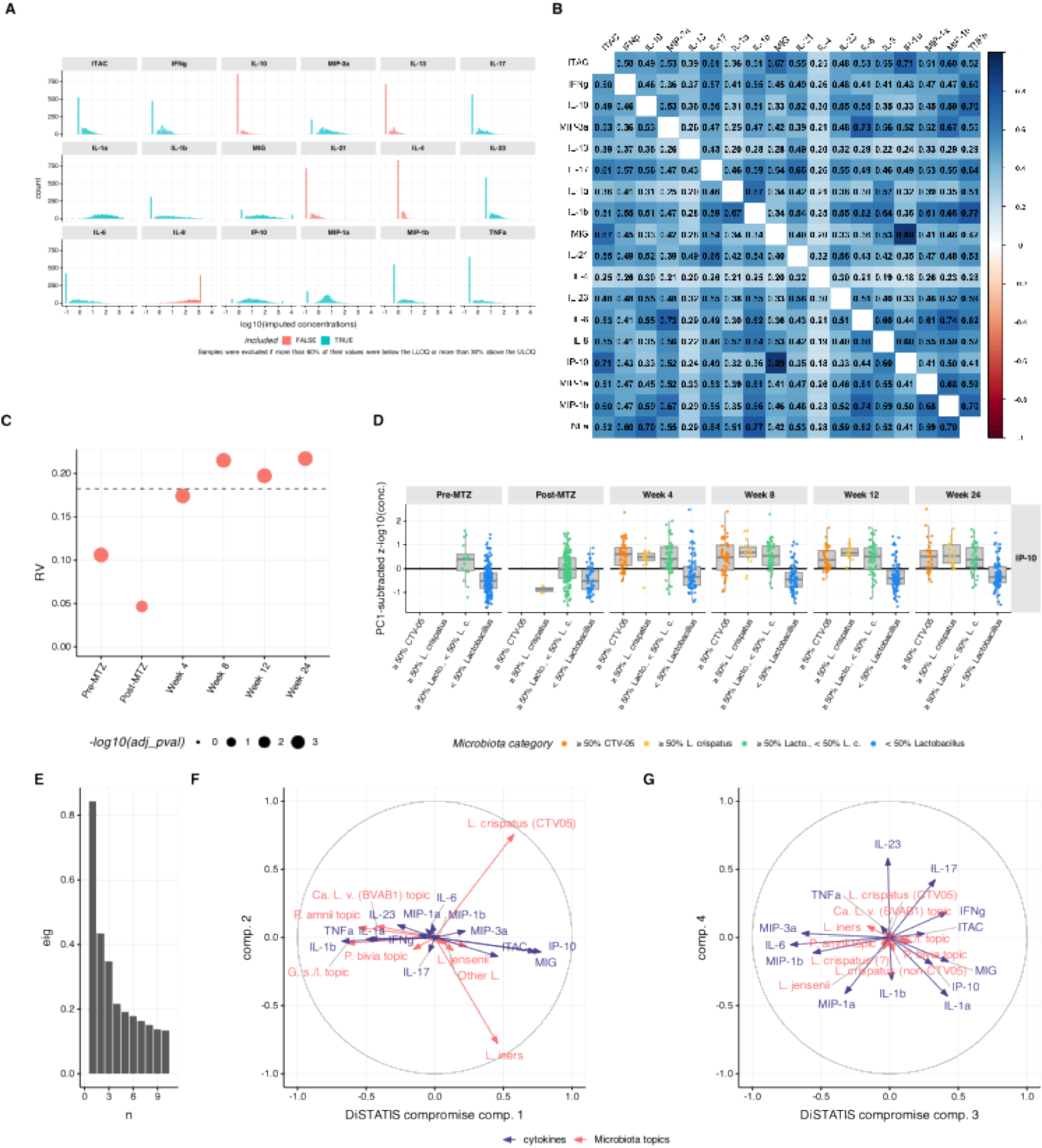
A. Distribution of unadjusted cytokine and chemokine concentrations within the dataset, with values below the lower limit of quantification (LLOQ) imputed at half the LLOQ and values above the upper limit of quantification (ULOQ) imputed at the ULOQ. Analytes were excluded from further analysis if >60% of values were below the LLOQ or >30% were above the ULOQ. B. Pair-wise Pearson correlation coefficients between log10-transformed concentrations of each cytokine. All correlations were significant (p < 0.05). C. RV coefficients (y-axis) between microbiota composition described in terms of topic proportions and cytokine/chemokine transformed concentrations at each visit (x-axis). Permutation test *p*-values were adjusted for multiple testing to control for the false discovery rate. The size of the dot is inversely proportional to the log_10_ of the associated adjusted p-value. The horizontal dashed line provides the value of the RV coefficient when computed on all visits combined. D. IP-10 adjusted log_10_(concentrations) (y-axis) by microbiota composition categories (x-axis) at each visit (horizontal panels). Microbiota categories are mutually exclusive such that the first category (≥ 50% CTV-05) encompasses samples in which the relative abundance of CTV-05 (within the overall microbiota) is larger or equal to 50%; the second category encompasses samples with ≥50% total *L. crispatus* but <50% CTV-05 relative abundance within the microbiota; the third category includes samples in which total *L. crispatus* relative abundance is <50%, but the total *Lactobacillus* relative abundance is ≥50%, and the remaining category includes samples with <50% *Lactobacillus*. Participants from both arms are included. E. DISTATIS screeplot from repeated DiSTATIS analysis, similar to analysis in Figure 4D but differentiating between CTV-05 and summed other *L. crispatus* strains. Eigenvalues (a.u.) of DISTATIS compromise for the first 10 latent components are shown. F. DISTATIS correlation circle for the analysis in **Figure S4E**, showing the 1st and 2nd latent components (similar to Figure 4E). G. DISTATIS correlation circle for the analysis in **Figure S4E**, showing the 3rd and 4th latent components (similar to Figure 4F).

**Figure S5.**
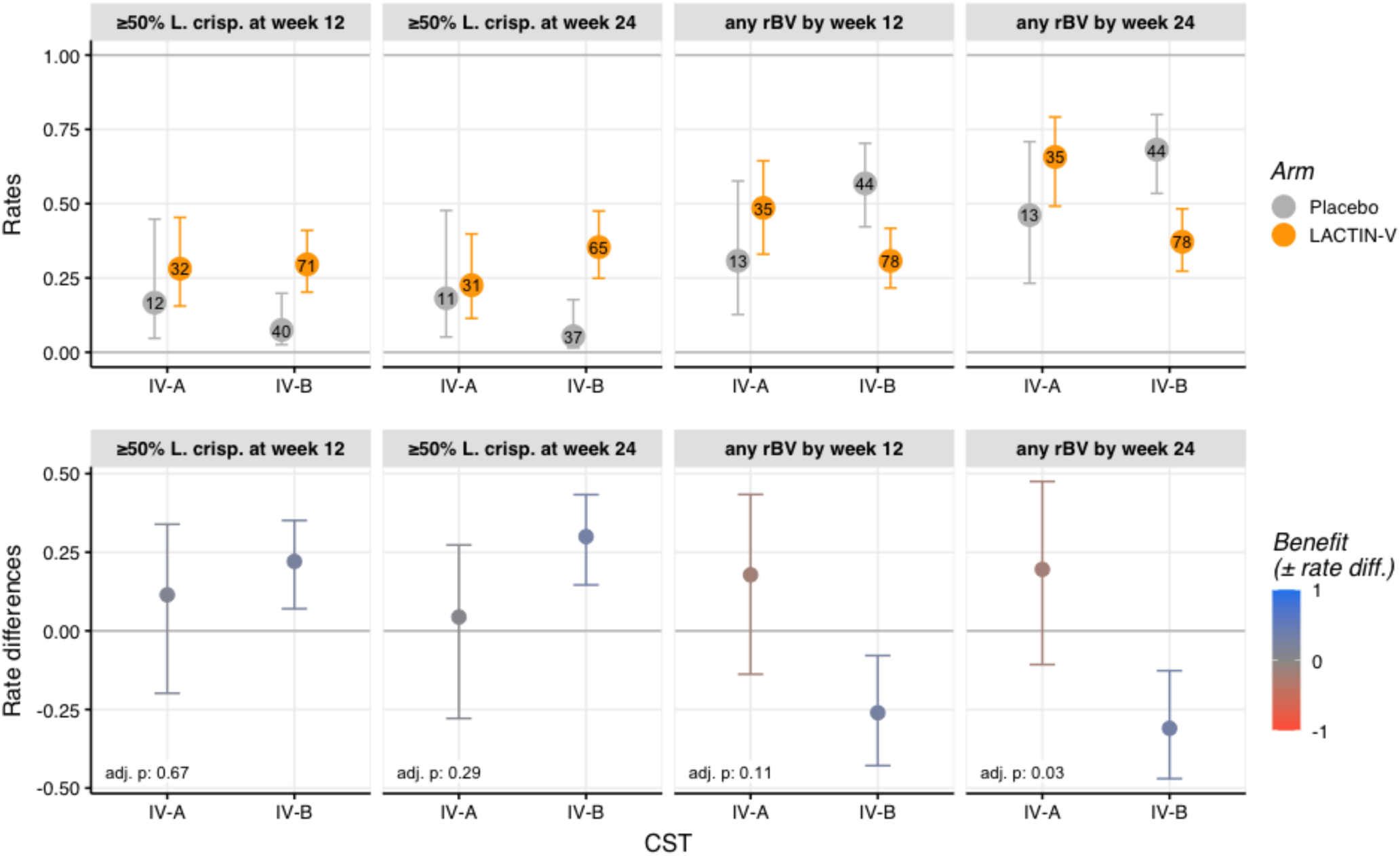
Rates and associated 95% CI of high-level *L. crispatus* colonization (≥50% of relative abundance) (y-axis) at week 12 or 24 (left two panels, top row) or rBV by Week 12 or 24 (right two panels, top row) by pre-MTZ CST in each arm (colors), and the corresponding rate differences and 95% CI between arms (bottom row), analyzed as in Figure 5C-F. Analysis is restricted to CSTs with at least two participants in each arm (*i.e.,* CSTs IV-A and IV-B). The color scale in the bottom row shows the degree of benefit (defined as the difference in rates between the two arms) in achieving *L. crispatus*-dominance (left panels) or reducing rBV (right panels), with blue indicating benefit with LBP treatment and red indicating benefit with placebo. *P*-values adjusted for multiple testing using the Benjamini-Hochberg correction of the analysis of deviance test for heterogeneity in treatment effects when stratifying participants by CST at the pre-MTZ visit. The centroid definition of CST IV-A includes *Ca*. Lachnocurva vaginae (BVAB1) as a predominant taxon while CST IV-B has higher abundance of *Gardnerella* and both share moderate *Prevotella* abundance (France et al., 2020).

**Figure S6.**
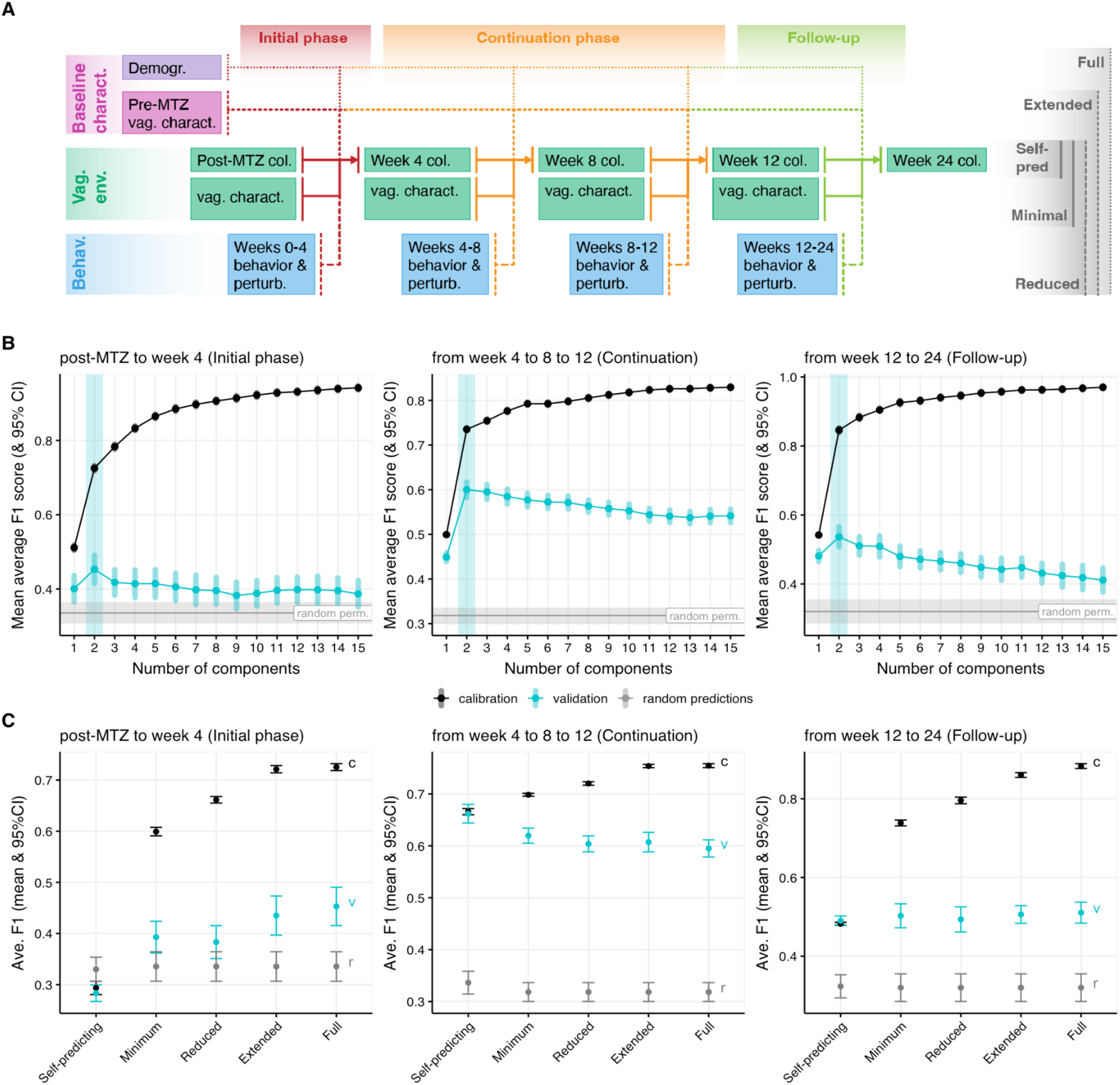
A. Diagram depicting MB-PLS-DA nested models. To assess whether blocks contributed to explaining colonization, we compared a series of nested models (far-right labels). The full model includes all blocks and variables, while the “self-predicting” model solely includes the microbiota categories at the previous visit. B. Number of latent component selection by cross-validation for the initial phase (left), continuation phase (middle), and follow-up phase models (S6A). Mean (dots) and 95% CI (vertical segment) of the average F1 score (y-axis) obtained when including k latent components (x-axis) on 20 random calibration (black) or validation (turquoise) set. For reference, values obtained for random permutations of the response labels are shown by the horizontal gray line and band. The number of selected latent components is shown by the light turquoise vertical band (here k = 2 for all phases). C. Contribution of nested models for the initial, continuation, and follow-up phase models. Mean and 95%CI of the average F1 score (y-axis) on the calibration (black, “c”), validation (turquoise, “v”), or for random predictions (gray, “r”) for each nested model (x-axis) including the number of latent components shown on the corresponding upper-panel.

**Figure S7:**
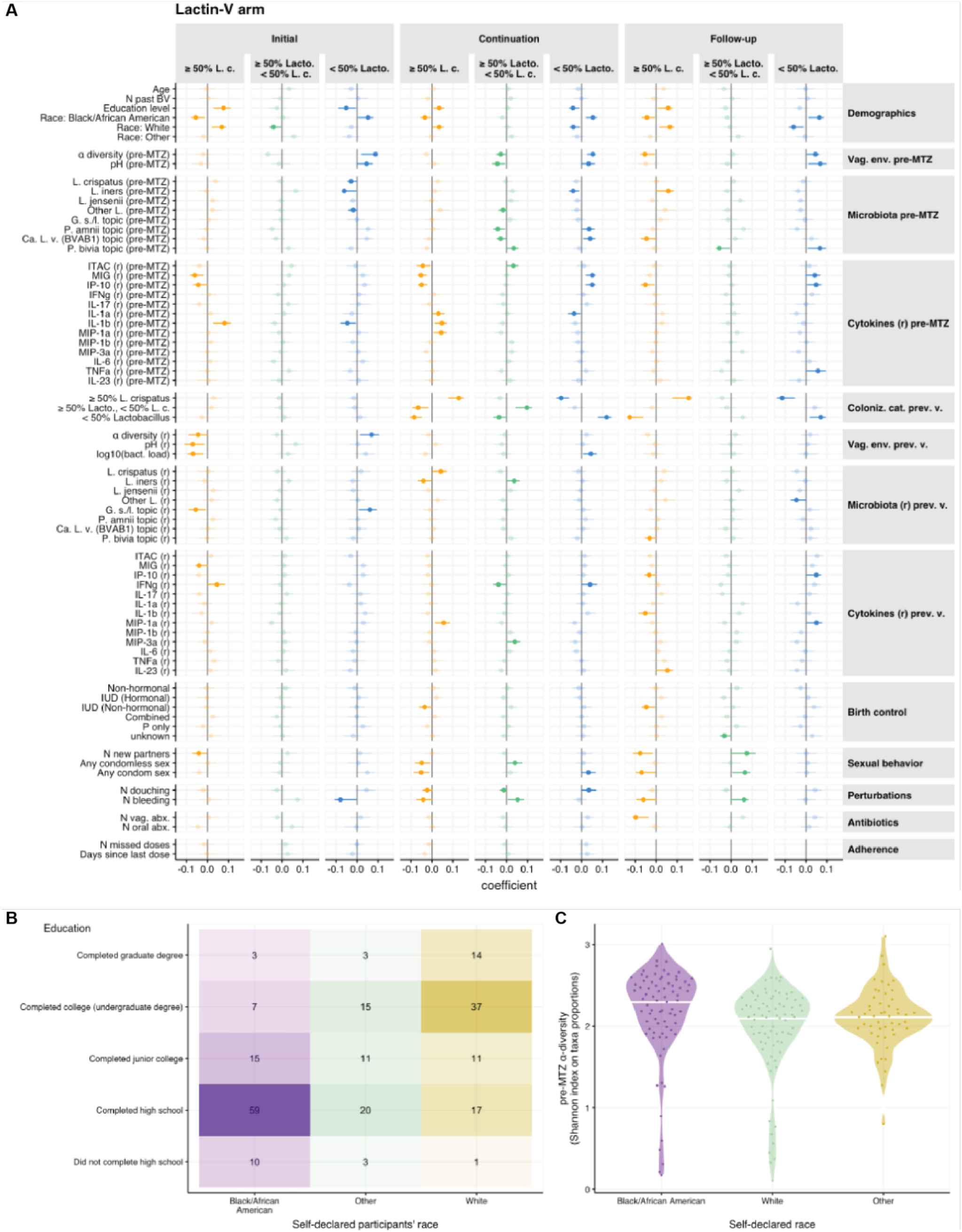
Variables associated with microbiota categories in LACTIN-V recipients. A. Coefficient of association (dots, x-axis) and associated bootstrapped 95% CI (horizontal segments) between explanatory variables (y-axis) and microbiota categories (colors, horizontal panels, see Figure 6A) for each phase of the trial (horizontal panels) for MB-PLS-DA models fitted on LBP recipient data (**Figure S6A**). Variables whose 95% CI include 0 are shown in transparent/lighter shades. Variables are shown grouped according to their assigned thematic blocks (Figure 6A, **Table S6**). B. Self-reported education levels are significantly associated with self-reported race (χ^2^ *p*-value < 0.01) C. Pre-MTZ α-diversity is higher in Black or African American participants than in White participants (*p*-value of *t*-test on association coefficient < 0.05).

**Figure S8:**
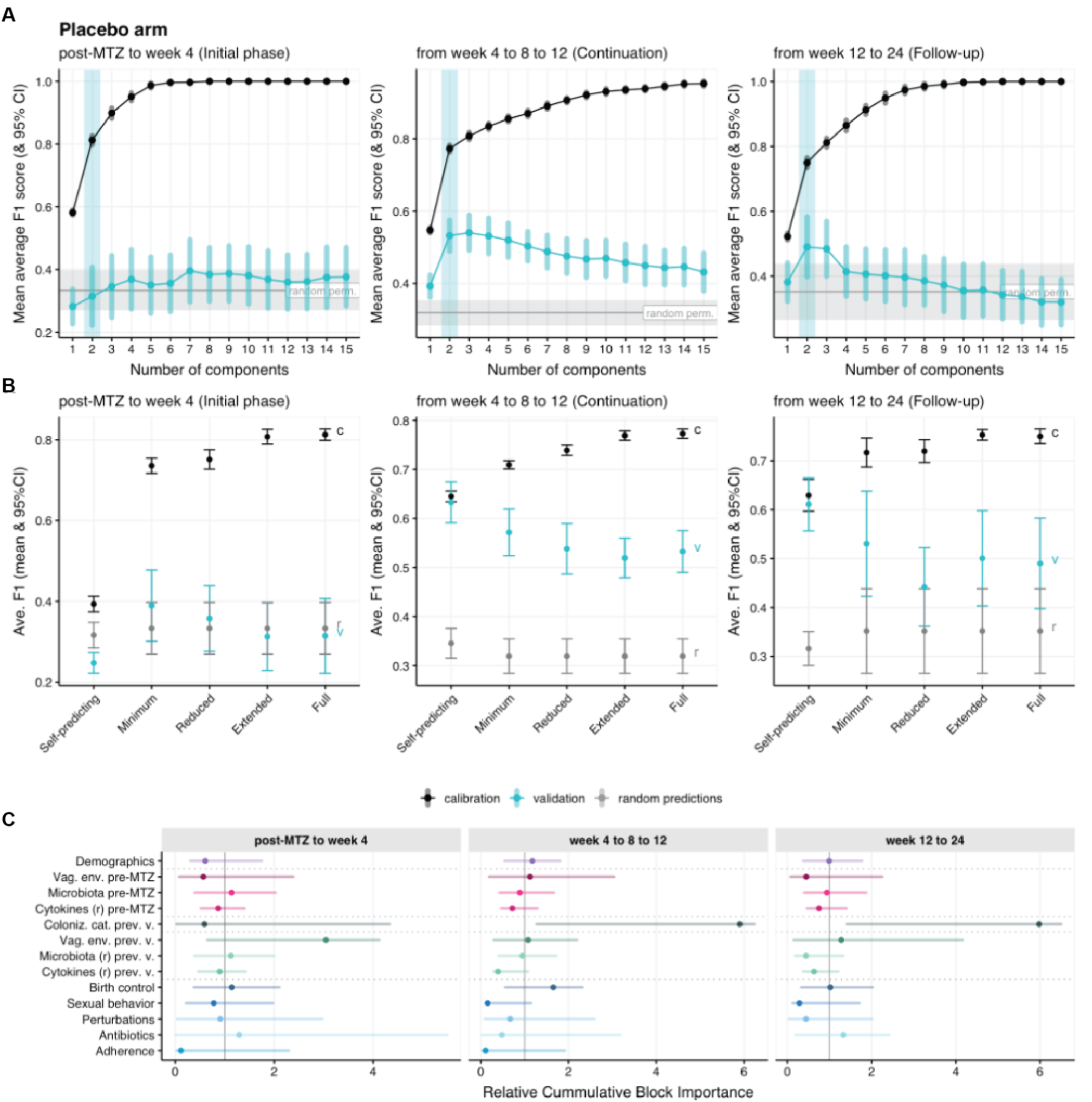
Multiblock analysis of factors associated with microbiota categories in placebo recipients. A. Number of latent component selection by cross-validation for the initial (left), continuation (middle), and follow-up phase (right) model fitted on the placebo arm data. Reads as **Figure S6B**. B. Contribution of nested models fitted on the placebo arm data for the initial (left), continuation (missing) and follow-up phase (right). Reads as **Figure S6C.** C. Relative cumulative importance indices in the placebo arm (x-axis, Methods) for each block (y-axis, color) for the initial phase model (post-MTZ to Week 4; left), continuation phase model (Week 4 to 8 to 12; middle), and follow-up phase model (Week 12 to 24; right), analogous to portrayal of LBP arm in Figure 6A.

**Figure S9:**
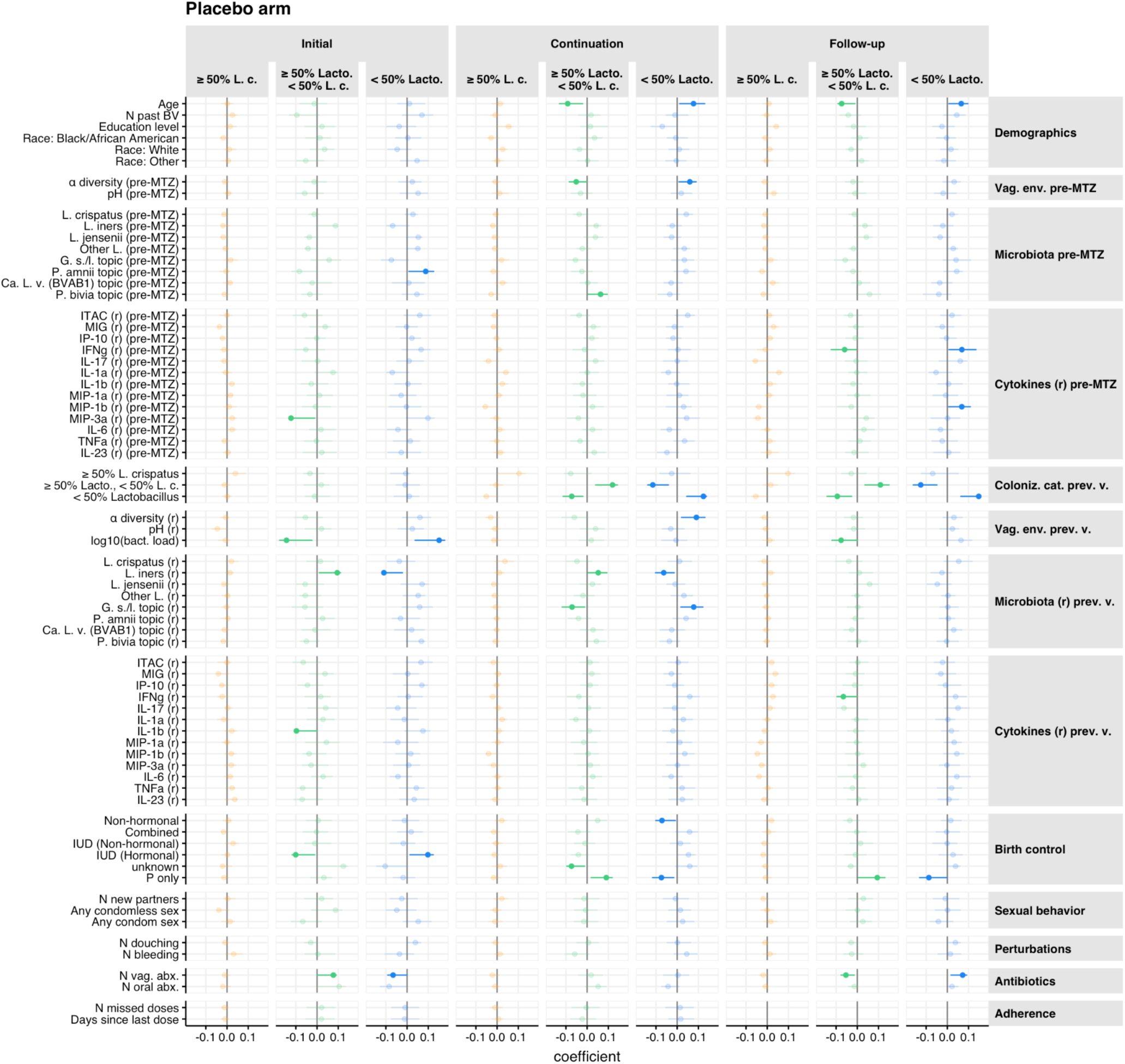
Variables associated with microbiota categories in placebo recipients Coefficient of association (dots, x-axis) and associated bootstrapped 95% CI (horizontal segments) between explanatory variables (y-axis) and microbiota categories (colors, horizontal panels) for each phase of the trial (horizontal panels) when MB-PLS-DA models are fitted on the placebo arm participants’ data. Variables whose 95% CI include 0 (suggesting no statistically significant associations) are shown in transparent/lighter shades. Variables are shown grouped according to their assigned thematic blocks (Figure 6A, **S8C Table S6**).

